# Inter-individual variability in lipoprotein proteomics reveals distinct patient clusters informative for disease pathogenesis and severity

**DOI:** 10.64898/2026.03.26.26349288

**Authors:** Maxime Nguyen, Soukaina Timouma, Hanyu Qin, Yuxin Mi, Charles Hinds, Stuart Mc Kechnie, GAinS Investigators, Thomas Gautier, Julian C. Knight

## Abstract

Lipoprotein composition is altered in sepsis, and supplementation with high-density lipoproteins has been reported to improve outcomes in experimental settings. In this study, we aimed to investigate the nature and inter-individual variability in the lipoprotein proteome to inform risk stratification and opportunities for precision medicine approaches. In a large proteomic dataset including 1134 patients (1781 samples) with sepsis and 149 healthy volunteers, we analysed 18 protein components of lipoproteins. We characterise heterogeneity of the lipoprotein proteome, defining three step-wise sub-phenotypes associated with increasing disease severity, one close to health, then an early phase patient group showing increased abundance of proteins that integrate HDL under inflammatory conditions (SAA1 and SAA2), then a group with decreased abundance of proteins that are components of HDL under healthy conditions that was associated with higher organ failure intensity (SOFA score) and increased mortality. We developed and externally validated a quantitative score reflective of lipoproteins alterations in sepsis, and machine learning predictive models to predict the LP class, advancing future individualised lipoproteins-based therapeutics in sepsis.

**Purpose:** During sepsis, the lipoprotein proteome is altered, impacting function and potentially driving severity. Many drugs targeting lipoprotein metabolism are available, but sepsis is a highly heterogenous disease, and there is no large-scaled exploration of lipoprotein alterations in sepsis. In this study, we aimed to explore lipoprotein proteome based sub-phenotypes and their implications in terms of immune dysfunction, organ failure, and mortality in sepsis. The secondary objective was to develop classifiers enabling risk stratification and personalised intervention for future trials.

**Methods:** In a large proteomic dataset, we identified 18 proteins that are components of lipoproteins. We conducted unsupervised clustering and compared those patient clusters in terms of immune function (SRS scores, differential gene expression analysis), organ dysfunction, and mortality. We then built a continuous score for lipoprotein alteration in sepsis. In the third part, we developed and externally validated two predictive machine learning models that can be used as classifiers (for sub-phenotypes assignment and continuous score calculation).

**Results:** We analysed data from 1134 patients (1781 samples) with sepsis and 149 healthy volunteers. On day 1 of ICU admission, we identified 3 clusters based on lipoproteins’ proteome profile (named LP3, LP2 and LP1). Transition between the cluster of lowest severity (LP3) to LP2 was characterised by an increased abundance in proteins that integrate HDL under inflammatory conditions (SAA1 and SAA2) and transition between LP2 and LP1 by a decreased abundance of a group of proteins that are component of HDL under healthy conditions. Those clusters were associated with sepsis response signature scores, organ failure, and mortality. Next, we developed a continuous score reflective of lipoproteins alterations in sepsis (LPq) and two machine learning classifiers to predict LP cluster membership and LPq. Analysis on an external dataset composed of 265 patients (353 samples) with COVID-19, sepsis and healthy volunteers enabled us to validate the robustness of our models and the external validity of our score.

**Conclusions:** We described the heterogeneity of lipoproteins’ proteome in sepsis and defined three sub-phenotypes of increasing severity. Those sub-phenotypes were mostly driven by the HDL proteome. Our results suggest that lipoprotein proteome alteration occurs as a continuum in patients with sepsis. The first step was characterised by increased abundance of proteins that integrate HDL composition under inflammatory conditions (SAA1 and SAA2), while the second step was characterised by decreased abundance of proteins that are components of HDL under healthy conditions. This second step was associated with higher organ failure intensity (SOFA score) and increased mortality. We developed and externally validated a quantitative score reflective of lipoproteins alterations in sepsis, and machine learning predictive models to predict the LP class, paving the way for individualised lipoproteins-based therapeutics in sepsis.

## Introduction

Sepsis is defined as a life-threatening organ dysfunction caused by a dysregulated host response to a pathogen^1^ and is responsible for high morbidity and mortality worldwide^2^. During sepsis, lipoprotein composition is altered, and supplementation with high-density lipoproteins has been reported to improve outcomes^3,4^, suggesting causality between lipoproteins alterations and adverse outcomes in sepsis. Lipoproteins are spherical structures synthesised by the liver and gut. They are composed of a hydrophobic lipidic core and a hydrophilic surface, associating amphipathic lipids and proteins. In addition to their role in lipid homeostasis, lipoproteins have also been demonstrated to have multiple functions including anti-inflammatory, antioxidant, anti-thrombotic, and endothelial protective effects that might be beneficial against sepsis^5,6^. Decreased cholesterol and increased triglyceride plasma levels are described in sepsis and associated with prognosis^7^. The protein component of lipoproteins is also altered by sepsis, which may result in dysfunctional particles^8^. Supplementation with some of those proteins, such as mimetic Apo-A1 (apolipoprotein A-I) peptide and PLTP (phospholipid transfer protein), has been demonstrated to improve outcomes in animal models^9–11^, suggesting that proteins composing lipoproteins are involved in the pathological septic process and therefore highlighting their therapeutic potential.

The history of drug discovery in sepsis is marked by numerous negative results, largely due to the heterogeneity of this clinical syndrome^12^. Understanding sepsis heterogeneity is crucial for effective risk stratification, therapeutic target identification, and individualised treatment. To address this issue, several sub-phenotypes derived from blood leukocyte transcriptomic profiles have been described, enabling a more precise characterisation of the heterogeneity of the host immune response during sepsis^13,14^.

We hypothesised that inter-individual differences in a patient’s lipoprotein proteome may contribute to sepsis immunopathology and clinical outcomes. Although observational studies have investigated the lipoprotein proteome in sepsis, either in comparison with healthy volunteers or according to survival status^6^, no large-scale study has yet explored the heterogeneity of the lipoprotein proteome in this condition. In this study, we aimed to explore interindividual variability in lipoprotein-associated proteins and their implications in terms of immune dysfunction, organ failure, and mortality in sepsis. The secondary objective was to develop predictive models enabling risk stratification and personalised intervention for future trials.

## Results

### Landscape of lipoprotein-associated proteins in sepsis

In order to investigate the nature and inter-individual variability in lipoprotein-associated proteins, we analysed a discovery cohort from the UK Genomic Advances in Sepsis study in whom we had generated proteomic data by mass spectrometry (LC-MS-MS)^15^. Data for 18 proteins which form an assembly with lipids (lipoproteins) were analysed following quality control from 1134 patients with sepsis (1781 samples: 572, 673, and 536 for day 1, day 3, and day 5, respectively) and 149 healthy controls. Among patients with sepsis, 520 (45.9%) were women, the median age was 65 years (interquartile range from 53 to 75 years), 743 (65.5%) had sepsis due to community-acquired pneumonia, and 391 (34.5%) due to peritonitis. The median SOFA score was 6 [4-9].

We first studied the abundance of the 18 proteins constitutive of lipoproteins identified in patients with sepsis, compared with healthy controls and changes over time. We found that sepsis (compared to healthy volunteers) was associated with an increased abundance of proteins described to integrate HDL composition during the acute phase, under inflammatory conditions (inflammatory HDL (i-HDL) proteins: SAA1, SAA2). Sepsis was also associated, to a lesser extent, with an increase of PLTP, APOF APOH and SAA4 abundance, with a decreased abundance of a group of proteins described as components of HDL in healthy conditions (healthy HDL (h-HDL) proteins: APOA1, APOA2, APOL1, APOM, PON1) and with a decreased abundance of APOA4, APOC4 and APOB (Figure 1A). Those differences globally appeared to be persistent on day 3 and day 5 (Figure 1B). i-HDL decreased over time, approaching the concentrations observed in healthy patients. Most h-HDL protein abundance also decreased over time, but this distances them further from the concentrations observed in healthy patients, suggesting a worsening of this component of proteome alteration (Supplementary Figure 1A).

**Figure 1.**
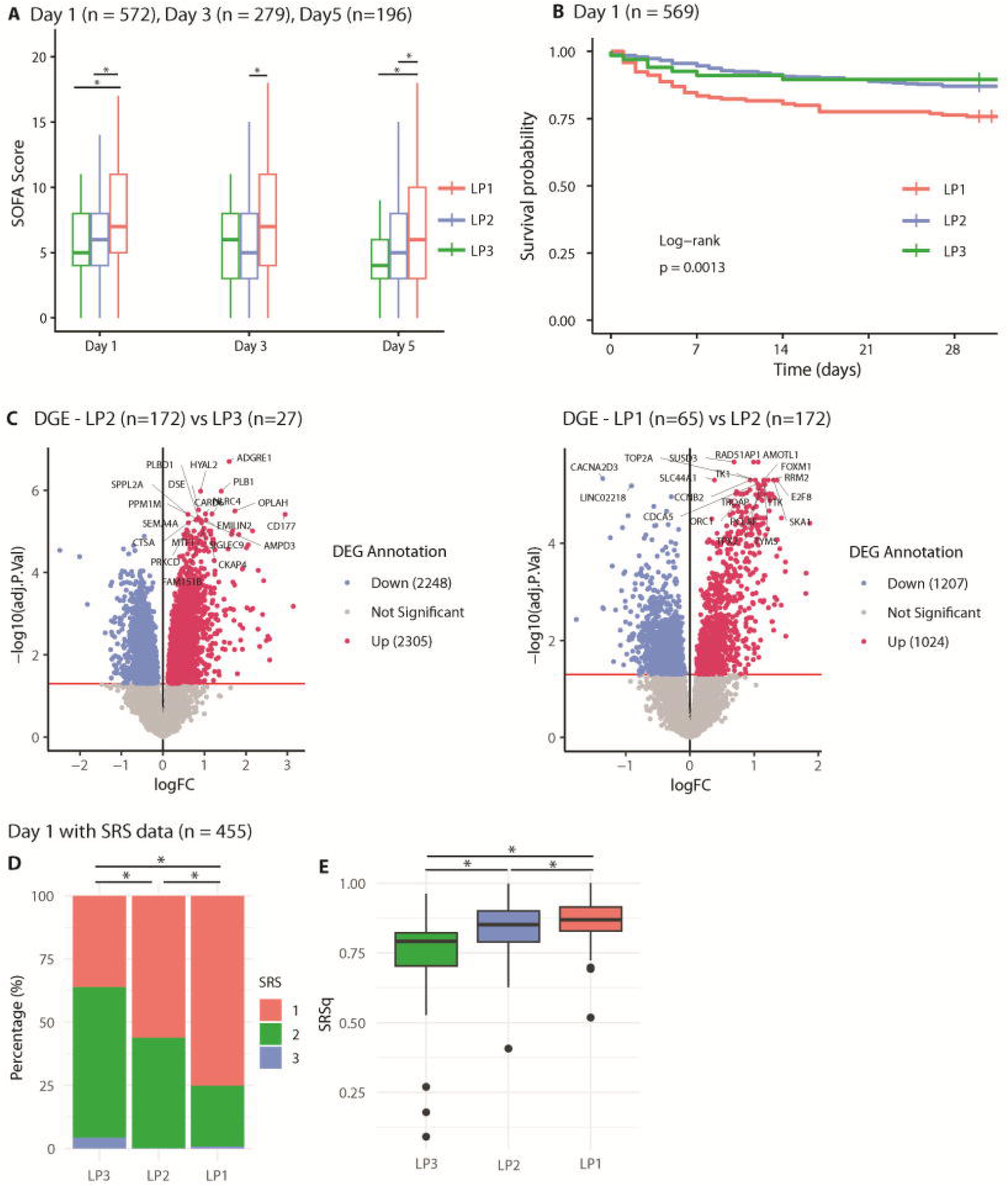
Lipoprotein-associated protein during sepsis on day 1 and longitudinal evolution. **(A-D)** Plasma proteomics data of sepsis patients, day 1 of admission (UK GAinS n=572 patients) (**A**) Volcano plot for differential lipoprotein assembly protein abundance between sepsis and healthy donors (n = 149) at day 1. Dashed line: adjusted p < 0.05; Red increased in sepsis; blue decreased in sepsis. (**B**) Heatmap displaying the longitudinal evolution (day 1, day 3 and day 5) of differential protein abundance between sepsis and healthy patients. Red increased in sepsis; blue decreased in sepsis. *: adjusted p < 0.05. (**C**) Volcano plot for differential protein abundance between ICU survivors and non-survivors. Dashed line: adjusted p < 0.05; Red increased in non-survivors; blue decreased in survivors. (**D**) Heatmap displaying the longitudinal evolution (day 1, day 3 and day 5) of differential protein abundance between survivors and non-survivors. Red increased in non-survivors; blue decreased in non-survivors. *: adjusted p < 0.05. ICU: Intensive Care Unit.

We then studied the association between the 18 proteins and sepsis severity by comparing patients that died in the ICU to patients that survived. We found that ICU mortality (compared with ICU survival) was predominantly associated with decreased abundance in h-HDL proteins (APOA1, APOA2, APOL1, APOC1, APOM, PON1). APOC4, SAA4 and SAA2 were of decreased abundance and PLTP was of increased abundance in patients that died (Figure 1C). Most associations between low h-HDL abundance and ICU mortality were persistent on day 3 but not on day 5 (Figure 1D).

Altogether, the HDL proteome (compared to other lipoproteins) was the most altered by sepsis. Consistent with previous literature, we found that sepsis was associated with an increase in a group of proteins described to integrate HDL composition under acute phase and inflammatory conditions (labelled i-HDL proteins) and a decrease in a group of protein described to be components of HDL under healthy conditions (labelled h-HDL proteins). On day 1, ICU mortality was mostly associated with a decreased abundance in h-HDL proteins but was not associated with increased abundance of i-HDL proteins. Our results also suggest that sepsis induced lipoproteins alteration is dynamic over time, prompting us to analyse day 1 samples separately.

### Clustering based on lipoprotein-associated proteins identified three sub-phenotypes

We next investigated variability in the host response and how this may aggregate between assayed proteins using principal component analysis (PCA). At baseline (day 1, n = 572), we found that principal Component 1 (PC1) correlated with h-HDL proteins (APOA1, APOA2, APOC1, APOL1, APOM, PON1), while Principal Component 2 (PC2) correlated with i-HDL proteins (SAA1 and SAA2) (Figure 2A). The correlation matrix confirmed the correlations between the h-HDL proteins and between the i-HDL proteins (Supplementary Figure 2A). PCA analysis and protein correlation conducted on the whole dataset (mixing the 3 time points) showed similar relationships between those variables, suggesting preserved patterns during the first 5 ICU days (Supplementary Figure 2B and C).

**Figure 2.**
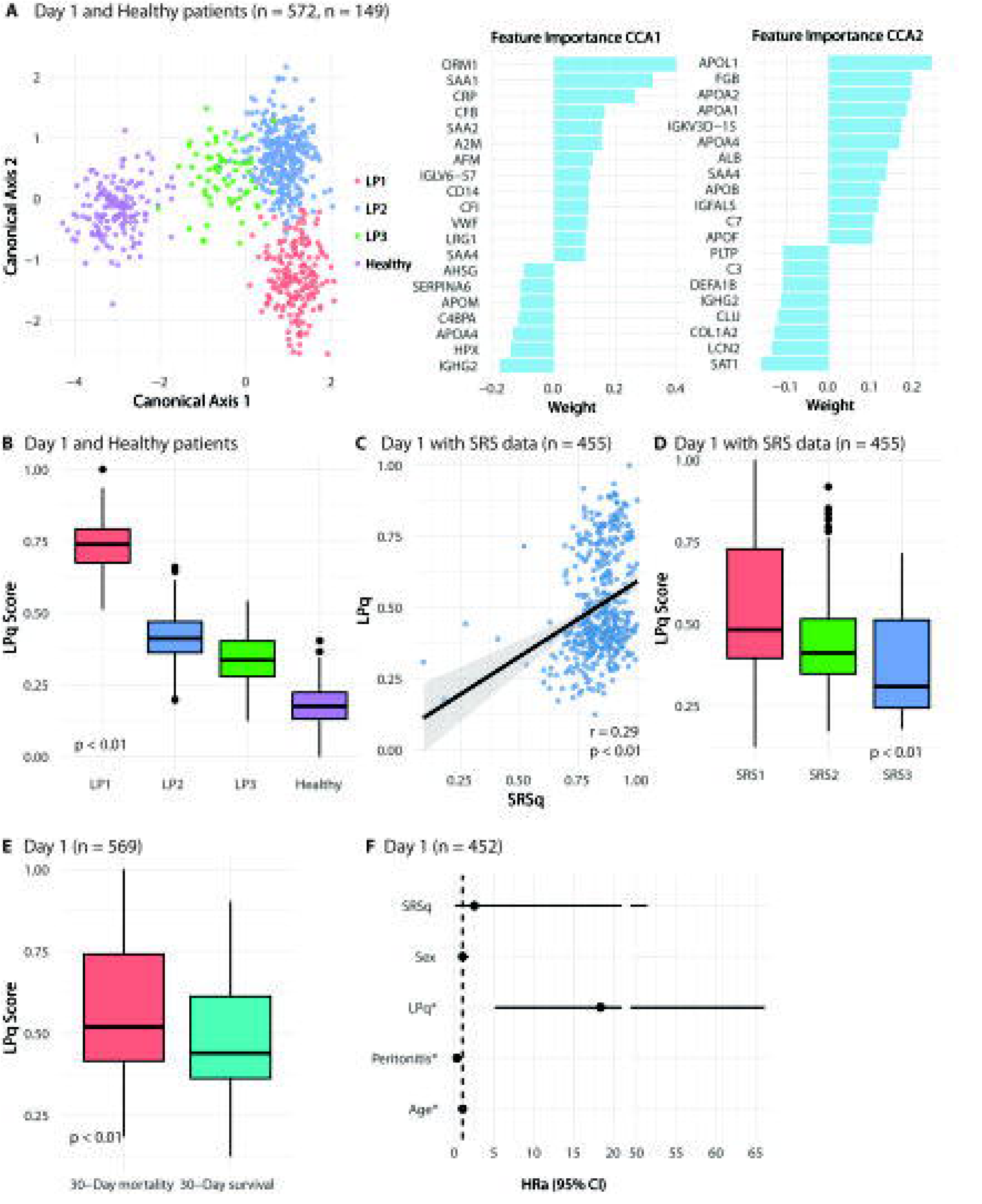
Lipoprotein-associated proteins’ clusters (LP) description. **(A-D)** Plasma proteomics data of sepsis patients, day 1 of admission (UK GAinS n=572 patients). **(A)** Principal component analysis, variable plot and proteins most associated with PC1 and PC2. Red: positive correlation; blue: negative correlation. **(B)** Principal component analysis individual plot. Colors represent LP clusters and dash line 95% ellipses. PCA was developed from Sepsis patients and healthy patients’ points (n=149) were secondarily projected. **(C)** Heatmap displaying individual protein abundance depending on LP cluster membership. **(D)** Volcano plots for differential protein abundance between LP3 and healthy patients (n = 149), LP2 and LP3, and LP1 and LP2. Dashed line: adjusted p-value < 0.05; Red: enriched; blue: decreased.

We then conducted three unsupervised clustering classifications (hierarchical clustering, K-mean clustering, and consensus clustering) using the 18 proteins of interest and the day 1 dataset. The optimal number of clusters was 3, based on the “NbClust” R package that determines the optimal number of clusters by summarising 36 metrics. Cluster memberships were consistent between the different techniques (74.3% of patients for hierarchical clustering and K-means, 83% between hierarchical and consensus clustering, and 82.2% between K-means and consensus clustering). Hierarchical clustering was retained as the best classifier for our data based on visual separation and 4 metrics (Silhouette, Dunn, Calinski Harabasz, Davies Bouldin) (Supplementary Material 1). Therefore, 3 clusters were selected. Lipoprotein cluster 1 (LP1) was found in 30.1% of patients (n=172), LP2 in 58.2% (333) of patients, and LP3 in 11.7% (67) patients.

After projection of the healthy controls (n=149) on the PCA graph, we observed that the individual graph showed a gradient separation between LP1, LP2, LP3 and healthy patients (Figure 2A and B). LP3 appeared as closest to health (Figure 2B). LP1 appeared as the most different from health with a shift toward lower PC1 (h-HDL) values, and LP2 seemed to represent an intermediate sub-phenotype mostly characterised by an increase in PC2 (i-HDL) values (Figure 2A). We then compared groups from healthy to most altered sub-phenotypes (healthy, LP3, LP2, and LP1) (Figure 2C and D). First, compared to healthy controls, LP3 mostly had an increased abundance of i-HDL proteins (SAA1, SAA2). LP3 also had, to a lesser extent, increased SAA4, PLTP, APOC1, APOH and APOF and decreased APOA1, APOA2, APOA4, PON1, APOC4, APOM and APOB abundances (Figure 2D). LP2 (compared to LP3) also mostly had an increased abundance in i-HDL proteins (increased SAA1, SAA2). To a smaller degree, LP2 (compared to LP3) had lower abundance of APOA1, APOA2, APOA4, PON1, APOC1, APOC2, APOC3 APOC4, APOM and APOE. Last, LP1 (compared to LP2) mostly had a lower abundance of h-HDL proteins under (APOA1, APOA2, APOC1, POL1, APOM, and PON1). LP1 also had a decreased SAA1, SAA2, SAA4, APOA4, APOC4 and increased APOH and PLTP (Figure 2C and D).

We last explored whether those differences in protein abundance described on day 1 were persistent on day 3 and day 5 (95 and 73 samples were available for LP1 on day 3 and 5 respectively, 151 and 97 for LP2, and 33 and 26 for LP3). Overall, the between-clustered differences reported at day 1 were same sided across time points. However, differences in i-HDL proteins between LP3 and LP2 seemed to decrease with time (Supplementary Figure 2D, Supplementary Figure 3A). On the contrary, differences in h-HDL reported between LP2 and LP1 seemed to be stable at day 3 and 5, suggesting persistent alterations (Supplementary Figure 2D, Supplementary Figure 3B).

Altogether, we describe 3 clusters based on lipoproteins proteome in sepsis. Evolution in protein content from healthy to the most altered sub-phenotype (LP1) shows that alterations seemed to occur gradually. The first step (from healthy to LP3 and from LP3 to LP2) was mostly characterised by an increased abundance in i-HDL proteins (SAA1 and SAA2) that appeared to regress with time. By contrast, the second step was mostly characterised by a decreased abundance in h-HDL proteins (APOA1, APOA2, APOC1, APOL1, APOM and PON1) and persisted over the first 5 days.

### Lipoprotein patient clusters are associated with sepsis response signature scores, organ failure and mortality

We then explored the clinical relevance of our clusters. Age, sex, and ethnicity were similar between clusters (Table 1). Peritonitis as a cause of sepsis had a higher prevalence in clusters with highest HDL proteome alterations (LP1 (55.8%), LP2 (28.8%), and LP3 (17.9%). On the contrary, community acquired pneumoniae had a higher prevalence in clusters closer to health (Table 1). LP1 had the highest clinical severity assessed by highest SOFA scores (Figure 3A); this difference seemed mainly driven by the cardiovascular and renal components of this score (Table 1). LP1 was also associated with higher mortality rates (Figure 3B).

**Figure 3.**
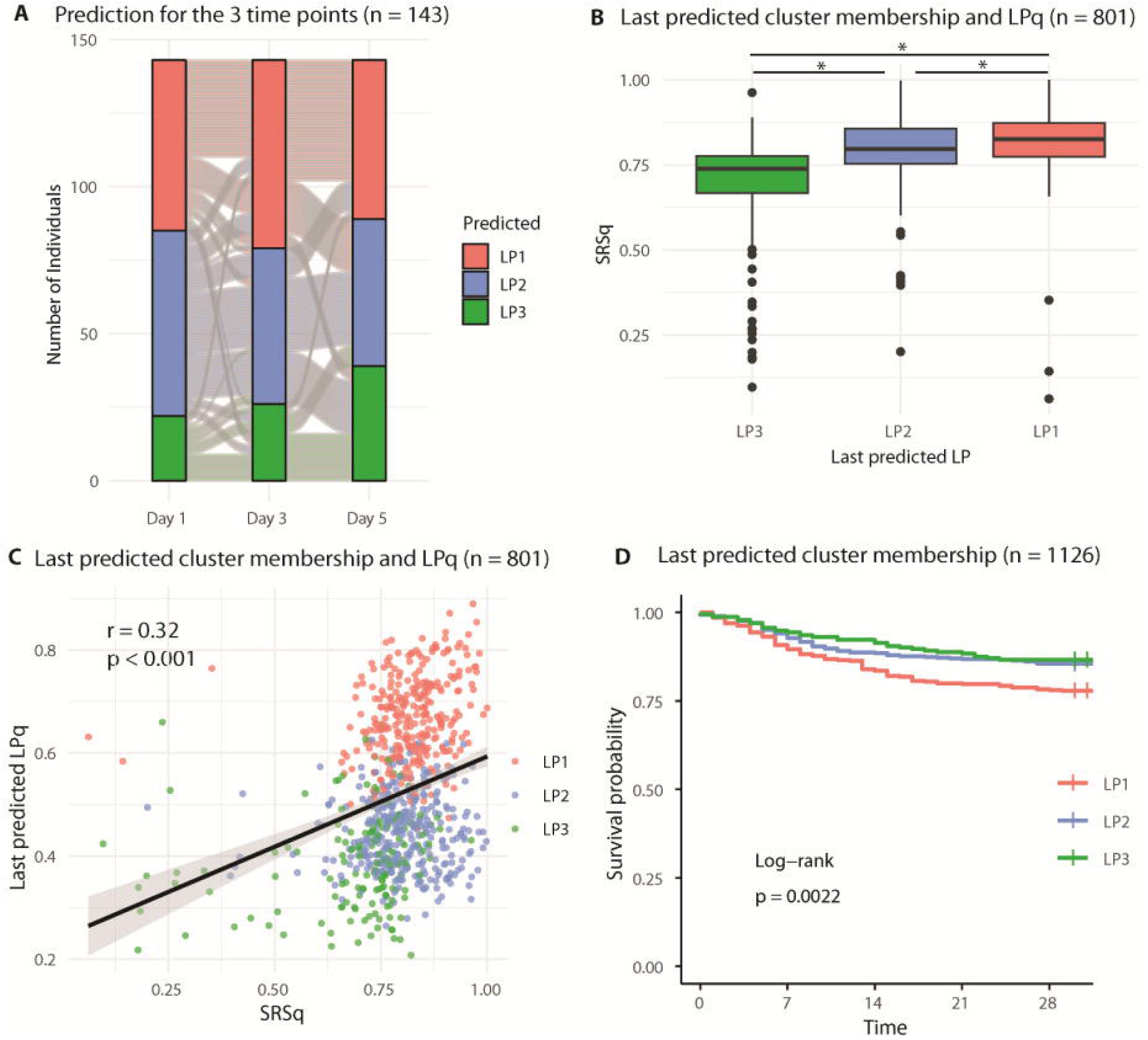
Lipoprotein-associated proteins’ clusters immune dysregulation and mortality. **(A-E)** Plasma proteomics and clinical data from UK GAinS. **(A)** Box plot for the association between LP cluster and SOFA score depending on the time of sampling. Green: LP3, Red: LP1, Blue: LP1. *: p < 0.05. **(B)** Kaplan-Meier plot for 30-day mortality depending on LP cluster membership. **(C)** Volcano plots for differential gene expression analysis between LP2 and LP3 and LP1 and LP2. Red line: statistical significance (adjusted p-value < 0.05). Red: upregulated; Blue: downregulated. **(D)** SRS category according to LP cluster membership. *: p < 0.05. **(E)** SRSq depending on LP cluster membership. *: p < 0.05. SOFA: sequential organ failure assessment; DGE: Differential gene expression; SRS: sepsis response signature.

**Table 1.**
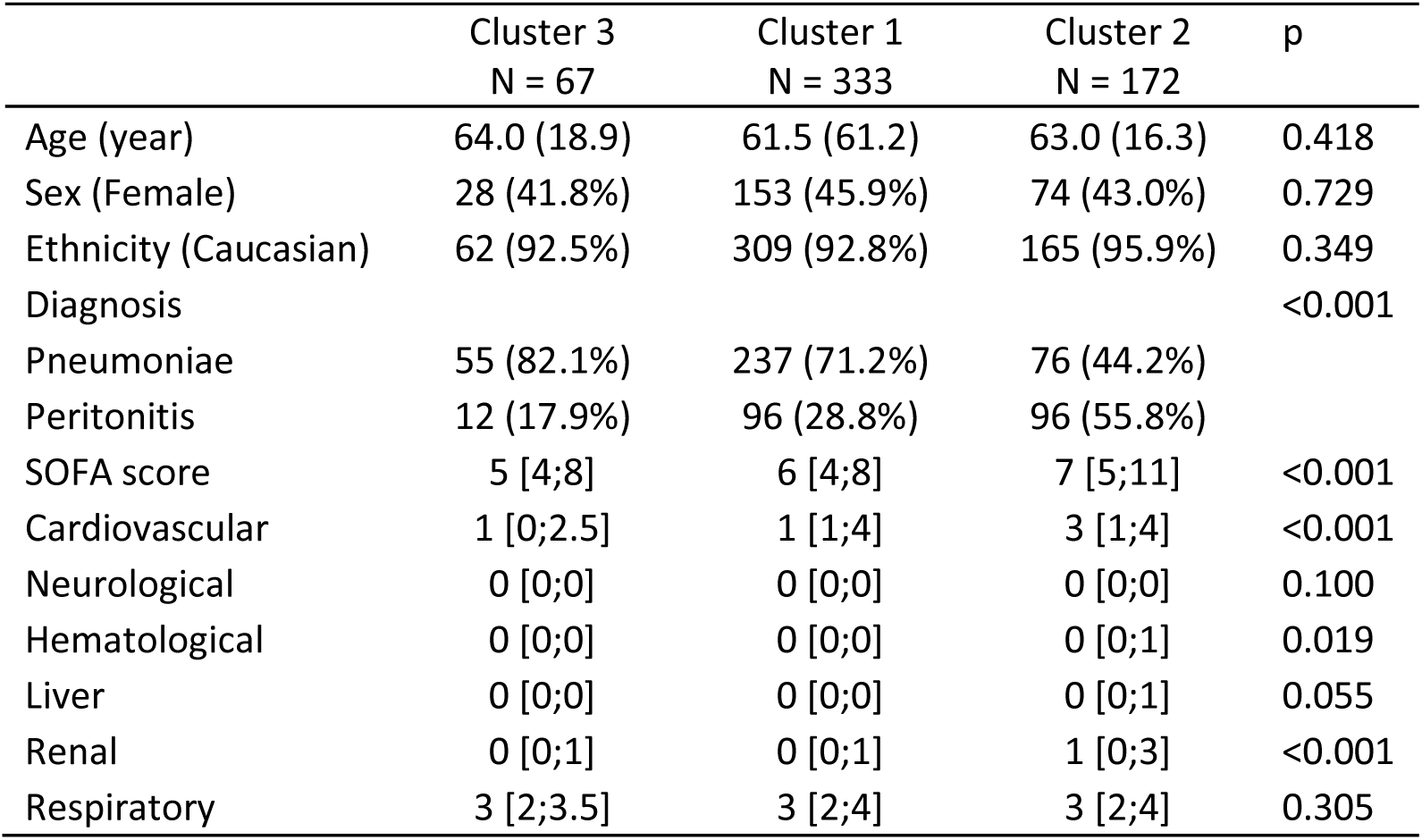
Baseline characteristics. SOFA: Sequential organ failure assessment;

|  | Cluster 3<br>N = 67 | Cluster 1<br>N = 333 | Cluster 2<br>N = 172 | p |
| --- | --- | --- | --- | --- |
| Age (year) | 64.0 (18.9) | 61.5 (61.2) | 63.0 (16.3) | 0.418 |
| Sex (Female) | 28 (41.8%) | 153 (45.9%) | 74 (43.0%) | 0.729 |
| Ethnicity (Caucasian) | 62 (92.5%) | 309 (92.8%) | 165 (95.9%) | 0.349 |
| Diagnosis |  |  |  | <0.001 |
| Pneumoniae | 55 (82.1%) | 237 (71.2%) | 76 (44.2%) |  |
| Peritonitis | 12 (17.9%) | 96 (28.8%) | 96 (55.8%) |  |
| SOFA score | 5 [4;8] | 6 [4;8] | 7 [5;11] | <0.001 |
| Cardiovascular | 1 [0;2.5] | 1 [1;4] | 3 [1;4] | <0.001 |
| Neurological | 0 [0;0] | 0 [0;0] | 0 [0;0] | 0.100 |
| Hematological | 0 [0;0] | 0 [0;0] | 0 [0;1] | 0.019 |
| Liver | 0 [0;0] | 0 [0;0] | 0 [0;1] | 0.055 |
| Renal | 0 [0;1] | 0 [0;1] | 1 [0;3] | <0.001 |
| Respiratory | 3 [2;3.5] | 3 [2;4] | 3 [2;4] | 0.305 |

Blood RNA sequencing data were available for 65, 172, and 27 patients (at day 1) in LP1, LP2, and LP3, respectively. Differential gene expression analysis revealed high numbers of differentially expressed genes (Figure 3C). Gene set enrichment analysis (GSEA) showed that LP2 (compared to LP3) had an upregulation of neutrophils and platelet-related pathways and downregulation of B-cell and complement-related pathways (Supplementary Figure 4A). LP1 (compared to LP2) had an upregulation of complement-related pathways and downregulation of T-cells and cytokine-related pathways (Supplementary Figure 4A). Analysis of the cytokine panel (n= 47 for LP1, n= 67 for LP2, and n = 13 for LP3) revealed globally increased cytokine levels in LP1 and decreased levels in LP3 (Supplementary Figure 4B).

Sepsis response signature (SRS) and SRSq scores derived from whole blood transcriptomic data capture aspects of the heterogeneity in the host’s immune response to sepsis. SRS1 (and high SRSq) is associated with sepsis severity and specifically identifies a sub-group of patients with myeloid dysfunction and immune suppression. SRS assignments were available in 455 patients (79.5%) on day 1. LP1 was associated with a higher proportion of SRS1 (Figure 3D). LP1 had the highest SRSq scores, followed by LP2 and LP3 (Figure 3E).

Altogether, LP clusters were associated with sepsis severity, mortality, and with the host’s response to sepsis, supporting their clinical relevance.

### A quantitative score for lipoprotein alteration in sepsis

We next sought to develop a quantitative score, reflective of lipoprotein alterations, for future patient risk stratification. We carried out a canonical correlation analysis on day 1 patients with sepsis patients and healthy controls (Figure 4A). This analysis was consistent with PCA dimensional reductions. The first 2 dimensions (Canonical correlation axis 1, CCA1, and canonical correlation axis 2, CCA2) enabled a good separation of LP clusters with the first canonical axis mostly correlated with i-HDL proteins and mostly separating healthy patients from LP3 and LP2 and the second canonical axis, mostly associated with h-HDL proteins, and mainly separating LP1 from other LP groups. We then weighed and used those 2 dimensions to create a continuous score named LPq. In view of its association with clinical severity and mortality, we chose to favor the discrimination of the LP1 cluster (CCA2 axis). The LPq score was highly associated with the LP cluster membership (Figure 4B). The correlation between LPq and SRSq was found to be weak (r = 0.29, p < 0.01) (Figure 4C), and both LPq and SRSq show associations with SRS^16^ (Figure 4D). This modest LPq/SRSq association is consistent with the fact that SRSq and LPq are derived from distinct molecular layers, transcriptomic and proteomic data respectively, suggesting that they capture partially non-overlapping aspects of disease severity. LPq was also associated with 30-day mortality (Figure 4E), independent of the main confounders identified (Figure 4F).

**Figure 4.**
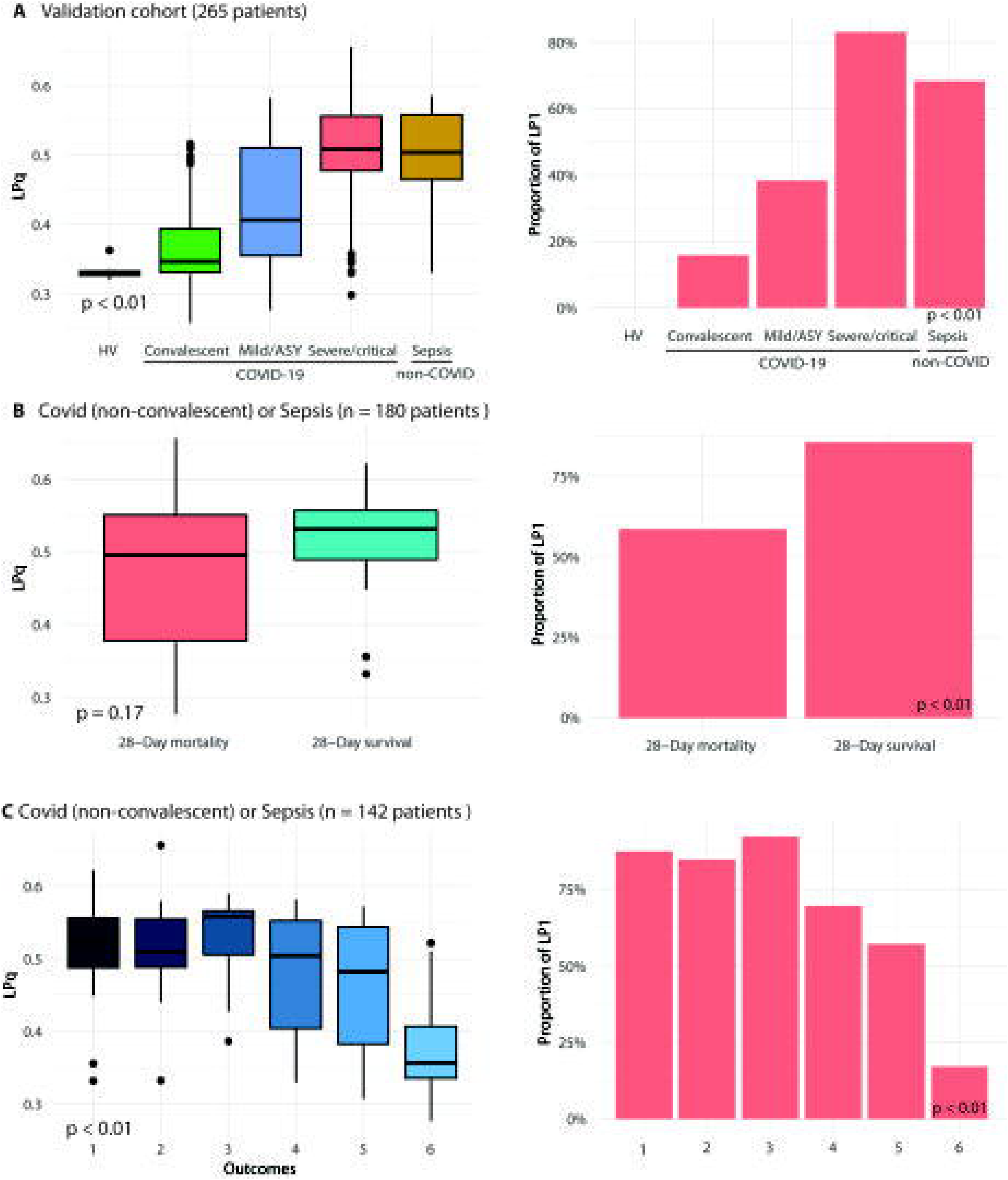
A quantitative score (LPq) for lipoproteins alteration in sepsis. **(A-F)** Plasma proteomics data from UK GAinS (Day 1 sepsis dataset, 572 patients and healthy volunteers, n = 149). **(A)** Canonical component analysis, individual map. Colors are LP clusters. The top 20 associated proteins are given for each component. **(B)** Boxplot representing LPq score depending on LP cluster membership. p is given for between groups comparisons. **(C)** LPq scores depending on SRSq scores. r is the spearman correlation coefficient. **(D)** LPq scores depending on the SRS group. p is given for between groups comparisons. **(E)** LPq score depending on 30-day mortality. p is given for between groups comparisons. **(F)** Cox models for 30-day mortality. Adjusted hazard ratios are represented along the x-axis and co-variate are represented along the y axis. *: p < 0.05. SRS: sepsis response signature; LPq: Lipoprotein quantitative score; CCA: Canonical correlation axis.

### Machine learning classifiers and regression model for lipoproteins-based stratification in sepsis

We trained random forest models on the Day 1 dataset (572 patients, 269 proteins). First, we built models to detect the importance of proteins that distinguish each class versus the other by including all proteins (269) available. The results were consistent with what we previously described, with APOA2 and APOL1, SAA1 and SAA2, being the top proteins separating LP1 from LP2, and LP2 from LP3, respectively (Supplementary Table 1). We next built a multiclass model and sought to minimise the number of proteins used in the model in order to facilitate its use (Supplementary Table 2). Good model performance was achieved with 10 proteins used as predictors comprising the 5 most predictive of CCA1 (ORM1, SAA1, CRP, IGHG2 and CFB) and the 5 most predictive of CCA2 (APOL1, FGB, APOA2, APOA1 and IGKV3D.15), and an optimal number of trees set to 550. AUCs were 0.91, 0.93 and 0.95 to predict LP3, LP2 and LP1 respectively (Supplementary Table 2, Supplementary Figure 5A), showing an overall good ability to distinguish between all classes. The overall accuracy was 84%, with an overall precision of 80%, a recall of 83%, and an F1-score of 81% (Supplementary Table 2). We also developed a regression model that enabled the prediction of LPq scores from those 10 proteins. For this model, R² was 0.77 and the agreement between predicted LP and LPq was good (Supplementary Figure 5B).

We further reduced the number of predictors to 5 for the multiclass model by keeping only CCA2 associated proteins (APOA1, APOL1, APOA2, FGB, and IGK3D.15). Models’ performances were slightly reduced compared to the 10 protein-based model, with an overall accuracy of 77%, precision of 68%, recall of 72%, and F1-score of 70% (optimal number of trees set to 670) (Supplementary Table 2). In particular, the discrimination between LP3 and LP2 that occurred mostly along the CCA1 axis was reduced (Supplementary Table 2, Supplementary Figure 5C and D).

### Longitudinal analysis on predicted LP and LPq suggests a continuum from LP3 to LP1

We then predicted LP membership and LPq score on the GAinS data set with proteomics data available (day 1, day 3 and day 5), based on the 10 protein predictive models (1134 patients, 1781 samples).

Among the 143 patients with data available at the 3 time points, we observed that between 2 consecutive time-points (day 1 to day 3 or day 3 to day 5), most patients had stable cluster membership (188 transitions, 65.7%) or transitioned to a cluster of adjacent severity (from LP1 to LP2, or LP2 and LP3, 71 transitions, 24.8%) (Figure 5A). Only a few patients had LP1-LP3 cluster transition (27 transitions, 9.4%), suggesting that clusters represent 3 successive states of increasing severity from LP3 to LP1 (Figure 5A).

**Figure 5.**
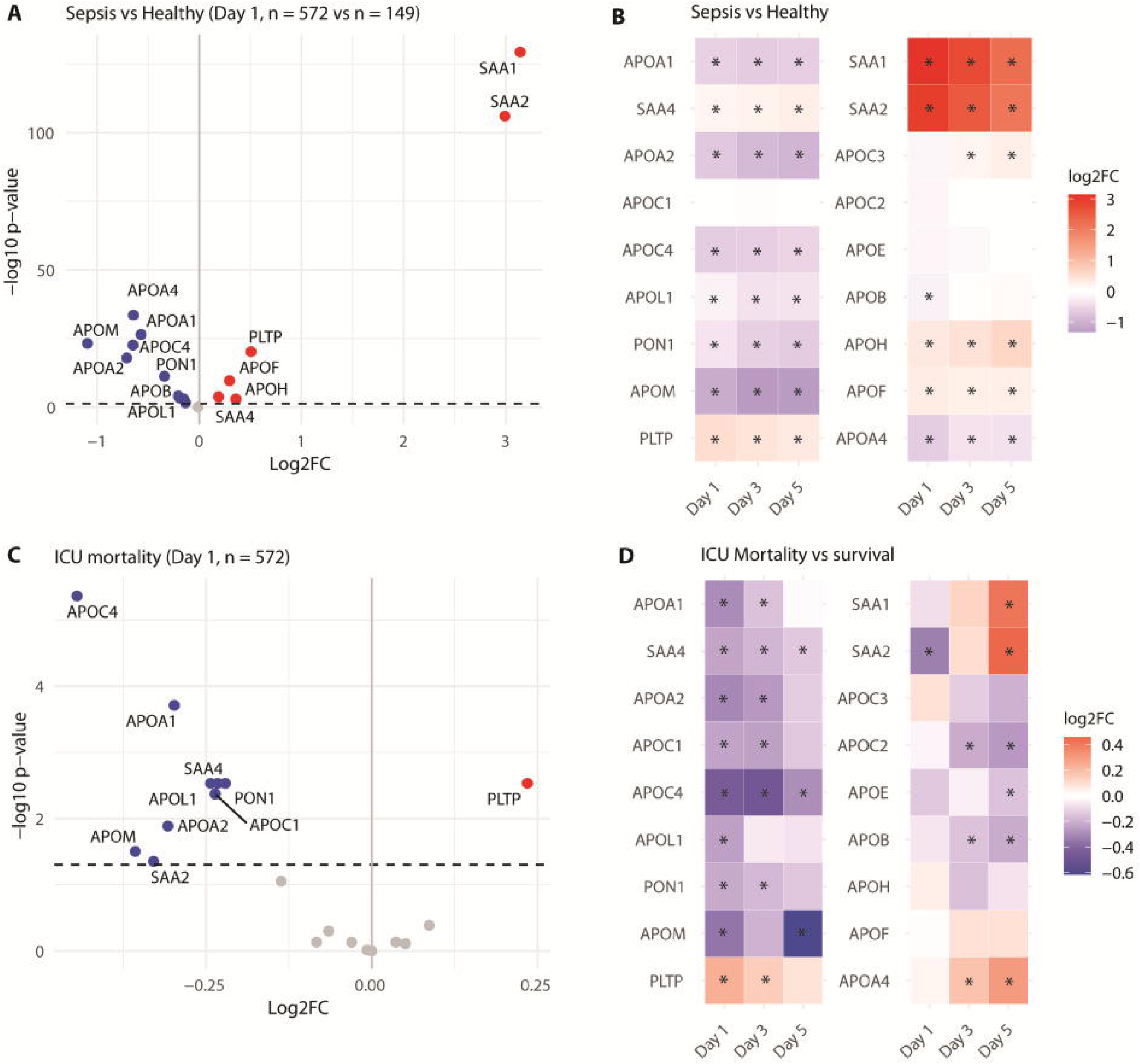
Longitudinal evolution of lipoprotein alteration. **(A-D)** Plasma proteomics data from UK GAinS (Sepsis dataset, all timepoints, 1134 patients, 1781 samples) **(A)** Alluvial plot representing predicted LP cluster membership evolution depending on the day of sampling (patients with sepsis and proteomic data at the 3 time points, n=143). **(B)** SRSq score depending on the last cluster membership. *: p < 0.05. **(C)** SRSq score depending on last predicted LPq. r is the Spearman correlation coefficient. **(D)** Kaplan-Meier plot representing 30-day mortality depending on the last LP membership. p is given for between group comparisons.

We then explored the clinical relevance of our score when predicted later in the disease time evolution. We used for every patient the last time point available (Day 1 (n=240), Day 3 (n = 358), Day 5 (n = 536)). Last cluster membership was associated with sepsis response signature scores (SRSq) (Figure 5B, C). Last predicted LPq was also associated with mortality (Figure 5D).

Altogether, LP and LPq prediction on the complete dataset demonstrate the robustness of our classifiers. Longitudinal analysis further supports that lipoprotein alterations occur as a continuum from LP3 to LP1. Those analyses support the relevance of our classification at later time points.

### External dataset validation

We last used an external dataset of patients with COVID-19 and non-COVID-19 sepsis from the COVID-19 Multi-Omics Blood ATlas (COMBAT) Consortium to validate our classifiers (COMBAT 2022). This dataset comprised 353 samples from 265 patients (105 patients hospitalised with COVID-19, 100 non-hospitalised patients with COVID-19, 22 healthy, 38 with non-COVID-19 sepsis). As not all proteins for the 10-protein based classifiers were available, we predicted LP and LPq using the 5 protein-based classifiers (using APOA1, APOL1, APOA2, FGB and IGK3D.15). In this cohort, LPq and LP were associated with diagnosis severity (Figure 6A). For the 82 samples (from 69 patients) with SRS data available, predicted LP and LPq were associated with SRSq but were not associated with SRS1 (Supplementary Figure 6A and B). In COVID-19 (non-convalescent) and sepsis patients, LP was associated with 28 days mortality, while LPq was not (Figure 6B). However, LP and LPq were associated with outcome severity (Figure 6C).

**Figure 6.**
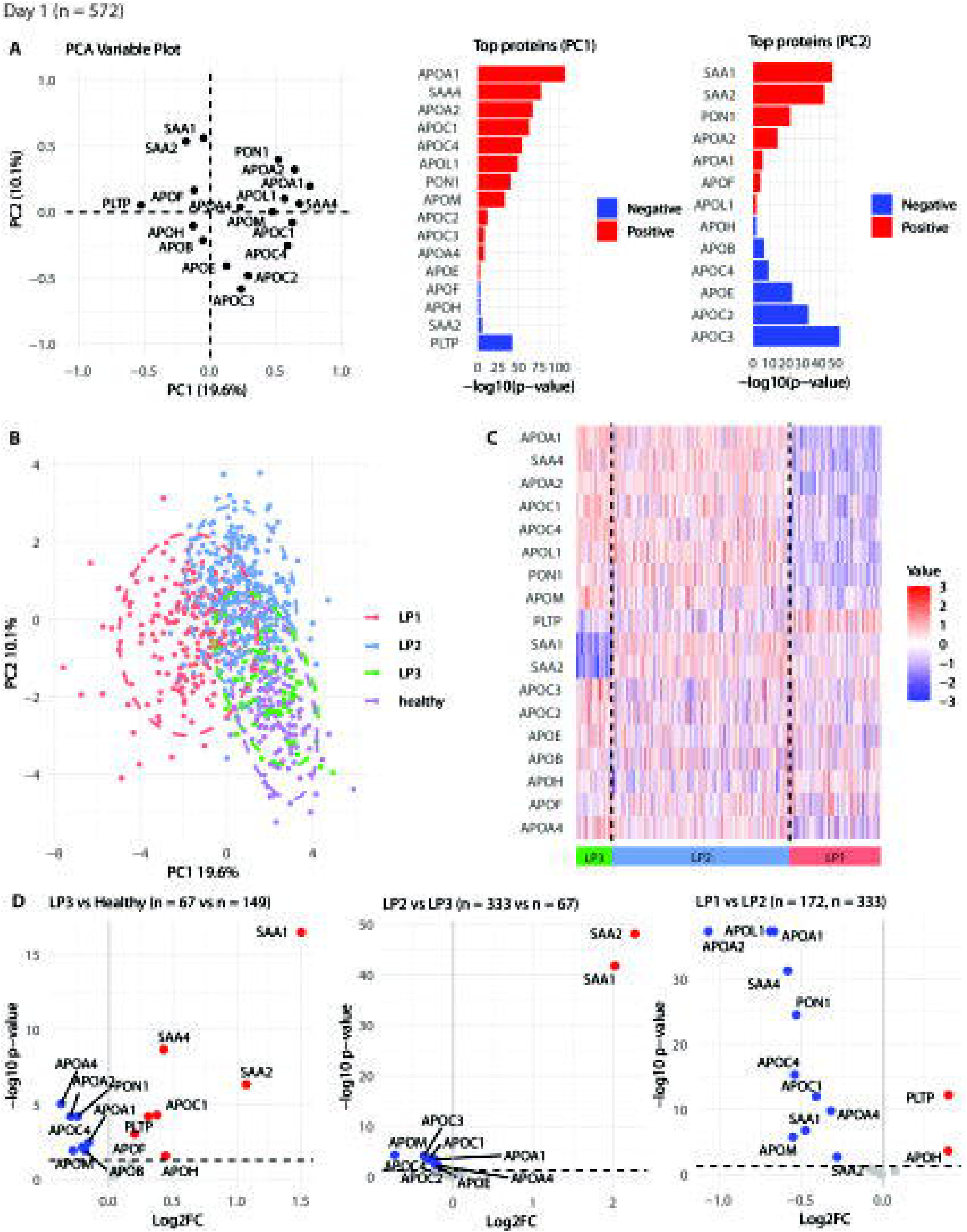
External cohort validation. **(A-C)** Plasma proteomics and clinical data from COMBAT (265 patients, 353 samples). **(A)** LPq and percentage of LP1 depending on disease severity. Convalescent, Mild/asymptomatic, severe refer to COVID-19 patients. p is given for between groups comparison. **(B)** LPq and percentage of LP1 depending on 28-Day mortality. p is given for between groups comparison. **(C)** LPq and percentage of LP1 depending on outcomes. 1: Death, 2: Intubated and ventilated, 3: non-invasive ventilation, 4: Hospitalised with O2, 5: Hospitalised, 6: Not hospitalised. p is given for between groups comparisons.

Altogether, the analysis of this external dataset supports the external validity of our classifiers and suggests their relevance in COVID patients.

## Discussion

In this paper, we describe the nature and variability of the lipoprotein proteome at an unprecedented resolution in patients with sepsis. Overall, key alterations were linked to HDL. Sepsis was predominantly associated with an increase in proteins previously reported to integrate HDL during the acute phase (i-HDL proteins, SAA1, SAA2), while ICU mortality was associated with a decrease in a group of proteins that are component of HDL under healthy conditions (h-HDL proteins, AOPA1, APOA2, APOC1, APOL1, APOM and PON1). Using the observed inter-individual variation in the sepsis lipoprotein proteome, we then described 3 sub-phenotypes associated with features of immune function. The first step of HDL proteome remodeling (between healthy volunteers and LP3 and between LP3 and LP2) was characterised by an increase in i-HDL proteins, while the second step (between LP2 and LP1) was characterised by decreased abundance of h-HDL proteins. This last sub-phenotype (LP1) was associated with higher clinical severity and mortality. We then developed a continuous score reflective of lipoprotein alterations in sepsis. Last, we developed and externally validated 2 machine learning classifiers to enable lipoprotein-based risk stratification in sepsis.

Sepsis induces modifications of both lipoprotein lipid content ^17,18^ and protein composition. In HDL, a switch from apolipoprotein-AI to SAA and modifications in enzymes with lower PON1, LCAT, CETP, and increased PLTP, endothelial lipase, and PLA2 are commonly described during sepsis ^5,8^. Our results are consistent with those previous findings, but the high sample size of our analysis enabled us to increase the resolution on this phenomenon. Whereas SAA has traditionally been described to replace APOA1 in inflammatory HDL ^7^ (and therefore the increase in SAA and the decrease in Apolipoproteins to occur simultaneously), our data suggests that the increase in i-HDL proteins and decrease in h-HDL proteins rather occurred as 2 successive steps in the lipoproteins’ alteration process. Furthermore, while the increase of SAA appeared to regress, the decrease in h-HDL proteins seemed persistent for the first 5 days. In line, LP1 was most associated with the intensity of organ failure and mortality.

In inflammatory states, HDLs are enriched in SAA, and SAA is often described as an acute-phase biomarker, like CRP. Aside from a chemotactic effect, SAA impact on immunity is not well described^19,20^ and whether SAA should be considered as a therapeutic target warrant further investigations.

The second step was mostly associated with a reduction in h-HDL proteins. Notably, this stage was not associated with a further increase in SAA protein abundance, underlying a different pattern of evolution. Several of those alterations are likely to influence the host’s response. APOA1 is the main protein component of HDL and plays a central role in HDL function. APOA1 enables HDL docking at cells’ surface and cholesterol efflux, which is a key mechanism of HDL anti-inflammatory effects. Indeed, cholesterol efflux modifies lipids raft composition which subsequently regulate immune receptor signaling such as T-cell receptors (TCR) B-cell receptors (BCR) Toll-like-receptor (TLR) and major histocompatibility complex II (MHCII)^21^. This is consistent with our findings that suggest altered TCR signaling in LP1 patients. Because administration of APO-A1 mimetics in animal models has been demonstrated to be protective against sepsis^10,11,22^, our results suggest APO-A1 as a potential therapeutic target in LP1 patients. APOM has a role in immunity through S1P metabolism and other h-HDL proteins, such as APOA2 and APOL1, and have also been described to have immune functions^23,24^. PON1 mediates HDL antioxidant effects upon LDL and may therefore be protective against sepsis. Altogether, those data suggest that the decrease in h-HDL lipoprotein may directly impact immune function, through the loss of HDL anti-inflammatory and antioxidant properties and therefore suggest those proteins to be promising therapeutic targets for LP1 patients.

Lipidome analysis reported association between lower cholesteryl ester and lysophospholipids and mortality at the whole blood^25^. At the cellular level, altered S1P metabolism was observed in immune cells^26^. Here, we did not have lipidomic data, and the joint analysis of HDL proteome and lipidome alterations could provide valuable additional insight.

There is increasing evidence of causality between lipoprotein alterations and adverse outcomes in sepsis. In animal models, several interventions targeting lipoprotein metabolism (such as recombinant HDL^4^ or recombinant PLTP^9^ administration) were demonstrated to improve survival. More recently, in a pilot study, administration of recombinant HDL to humans with sepsis was associated with reduced markers of inflammation^3^ and CETP inhibition has been shown to improve survival during sepsis by maintaining high HDL levels ^27^. Nevertheless, many therapeutics targeting immunity that had demonstrated experimental efficacy have failed to demonstrate efficacy in sepsis, most likely due to the disease’s heterogeneity. Therefore, our results, by providing a comprehensive approach to lipoproteins alterations in sepsis, appear as an essential first step, paving the way for individualised therapeutic interventions centered on lipoproteins. The progressive evolution of lipoprotein alterations is of particular interest in this process as it may enable optimizing the timing of interventions.

Our study has several limitations. The data are observational, and no causality can be inferred. Our proteomic dataset was derived from whole blood samples, and proteomic measurement of specific lipoprotein plasma fractions could have increased the resolution of our analysis. We did not have data on the lipid composition of lipoproteins, which would have helped further interpret our findings. Due to incomplete data, some analyses, in particular cytokine analysis, have a low sample size and may therefore lack power.

## Conclusions

We described the heterogeneity of lipoproteins proteome in sepsis and defined 3 sub-phenotypes. Those sub-phenotypes were mostly driven by HDL proteome alterations and were associated with the host’s response to infection, clinical severity, and mortality. We then developed a quantitative score reflective of lipoproteins alterations in sepsis. Last, we developed and externally validated 2 classifiers and a regression model enabling LP and LPq prediction from 5 proteins, paving the way for lipoprotein-based personalised treatment in sepsis.

## Material and methods

### Ethics

This study consisted of a retrospective analysis of previously published prospective cohorts^15,28,29^. Ethics approval was granted nationally and locally for individual participating centers, and we obtained informed consent from the patient or their legal representative (Scotland A Research Ethics Committee reference number 05/MRE00/38, Berkshire Research Ethics Committee (08/H0505/78), South Central Oxford REC C (19/SC/0296 and 06/Q1605/55).

### Patients

Data from the GAinS cohort with proteomic analysis are presented in this manuscript. Patients from the GAinS cohort were included in 34 UK intensive care units (ICU) between November 2006 and May 2018. Patients’ blood was sampled on day 1 and/or day 3 and/or day 5 of ICU admission. This study enrolled adults (> 18 years) with sepsis due to community-acquired pneumonia or fecal peritonitis (diagnosis based on international consensus criteria^30^). Healthy volunteers were also included in this cohort. Additionally, we used a dataset of patients from the COMBAT study ^28^. This data set included patients with confirmed COVID-19 of various severity, (recruited between 13 March and 28 April 2020), sepsis, and healthy patients.

Inclusion criteria were: patients admitted to Oxford University Hospitals NHS Foundation Trust, with a syndrome consistent with COVID-19 and a positive test for SARS-CoV-2 using reverse transcriptase polymerase chain reaction (RT-PCR) from an upper respiratory tract (nose/throat) swab.

Exclusion criteria were: Patients < 18 years old or with active malignancy or receiving significant immunosuppression (greater than an equivalent of 40mg once a day of prednisolone) prior to admission, or with a clear alternative cause for symptoms and hospital presentation.

Additionally, we used a dataset of patients from the COMBAT study^28^. This data set included patients with confirmed COVID-19 (recruited between 13 March and 28 April 2020) of various severity, sepsis, and healthy patients.

The patients included were admitted to Oxford University Hospitals NHS Foundation Trust, with a syndrome consistent with COVID-19 and a positive test for SARS-CoV-2 using reverse transcriptase polymerase chain reaction (RT-PCR) from an upper respiratory tract (nose/throat) swab.

The patients excluded were < 18 years old or with active malignancy or receiving significant immunosuppression (greater than an equivalent of 40mg once a day of prednisolone) prior to admission, or with a clear alternative cause for symptoms and hospital presentation.

### Proteomics

In the GAinS cohort, 269 proteins were measured by LC-MS-MS. We used data from the clean normalised count matrix previously published ^15^. Previous clustering analysis was also used. This previous analysis described 3 plasma protein-based clusters (SPC1, SPC2, and SPC3), with SPC1 displaying the most severe outcome. In the COMBAT dataset, 105 proteins were measured by mass spectrometry (LC-MS-MS) in 257 patients (340 samples). We used data from the clean normalised count matrix previously published^28^.

### Other Assays

Previously generated bulk RNA-seq datasets were used. Method and data pre-processing were described in previous papers ^29^. Sixty-five cytokines were measured in selected patients, using the ProcartaPlex Luminex platform (ThermoFisher Scientific) ^15^.

### SRS

SRS class was initially determined by unsupervised clustering methods from the whole blood transcriptome and can be predicted using either a 7-gene or 19-gene signature. SRS1 indicates higher immune dysfunction compared to SRS2 ^13^. The SRSq score is a quantitative score of immune dysregulations developed from the SRS classification that enables sepsis-induced immune dysregulation as a continuum from health to highly altered states^16^

### Differential protein abundance analysis

Differential protein abundance analysis was carried out using the package “limma” on R.4.4.1 (R version). P-values were corrected for repeated testing using the Benjamini-Hochberg method. Protein correlations were calculated using Pearson’s method. Because a patient could have several time points of sampling, time evolution was evaluated by mixed linear modeling (random intercept). Protein abundance was the dependent variable; time was used as a fixed factor and patient identifier as random effect.

### Patient clustering based on lipoprotein profile

Eighteen proteins constitutive of lipoproteins were available in the dataset and were used for clustering. Clustering was carried out upon data from day 1. Principal component analysis (PCA) was applied to the dataset (using the R package “FactomineR”), and 11 dimensions were kept, retaining more than 80% of the variance (based on scree plot analysis). Hierarchical clustering, K-means clustering, and consensual clustering were carried out. The optimal number of clusters was determined using the “NbClust” R package for hierarchical and K-means clustering. This package summarises 36 metrics to determine the optimal number of clusters. Hierarchical clustering was carried out using Euclidean distance and Ward’s method (“dist” and “hclust” functions). Consensus clustering was carried out with the “ConsensusClusterPlus” R package, using 1000 iterations of hierarchical clustering. The proportion of items to sample was set to 80% and the proportion of features to sample to 95%. The area under the cumulative distribution function curve was analysed to determine the optimal number of clusters. The final clustering method was chosen based on the visual separation on PCA and 4 metrics (Silhouette, Dunn, Calinski Harabasz, Davies Bouldin) (Supplementary Material 1).

### Clinical data analysis

Quantitative data are presented as median [Q1; Q3] or mean ± standard deviation, depending on their distribution. Missing data was considered to be random and was omitted. Clinical data were compared using the Student t-test, Kruskal-Wallis non-parametric test, chi-square test, or the Fisher exact test as appropriate (“CompareGroups” R package). Correlations were calculated using Spearman method. SRS and SRSq were calculated using the SepstratifieR R package ^16^. A multivariate linear regression was carried out to explore whether cluster membership was associated with SRSq independent of the main confounders identified by the bivariate analysis. SRSq was transformed (SRSq^3) to improve residual normality. The normality of the residual was checked graphically. Thirty-day mortality was analysed (“Survival” R package), data were represented on Kaplan-Meier plots (“Survminer” R package) and compared using the log-rank test. Cox models were used to adjust survival analysis on potential confounders.

### Bulk RNAseq analysis

For Bulk RNA-seq processing, gene expressions related to X and Y chromosomes were filtered out. One sample was identified as an outlier by PCA, and one sample with less than 1 million reads were excluded. Genes were filtered using the “filterByExpr” function of “edgeR” R package with default parameters. Differential gene expression (DGE) depending on clusters was carried out using “limma” R package. After differential gene expression, GSEA was carried out using the GSEA function (“clusterProfiler” R package), gene were ranked by log-fold change (log-FC) values, gene sets were selected from “msigbdr” (Reactome) and number of permutations was set to 1000. P-values were adjusted using the Benjamini-Hochberg method. Among significant pathways (q-value < 0.05), the 10 with higher NES and the 10 with lowest NES were plotted.

### Random forest classification models

Random Forest classifiers were implemented in a Python 3.11 environment using the “sklearn” package to perform supervised classification of LP classes using the protein abundance data. A python module, “RFmodule v. 0.1.0” (https://pypi.org/project/RFmodule/) was developed to perform hyperparameter optimisation as well as to build and train models.

#### Hyperparameter optimisation via Randomised Search

To identify optimal hyperparameters for the Random Forest classifiers, we employed a randomised search strategy using cross-validated grid exploration. This approach efficiently samples a broad range of parameter combinations to maximise model performance while controlling computational cost. The dataset was split into training and testing subsets using a stratified random split, with a predefined proportion allocated to testing. This ensured unbiased model evaluation and reproducibility through a fixed random seed. The search space for hyperparameters included:

- Number of trees: ranged from 5 to 1500 trees, in increments of 5.
- Maximum tree depth: integers from 1 up to a value informed by the logarithm of the number of features plus a buffer to allow for deeper trees.
- Minimum samples required to split a node: varied from small values (2) up to large fractions of the training sample size to control node splitting granularity.
- Minimum samples required at a leaf node: similar range to minimum samples required to split a node, including small and large leaf sizes to prevent overfitting or underfitting.
- Number of features considered for splitting: multiple values including square root, log2, fractions of total features, and small integers were tested to optimise feature subset size per split.
- Bootstrap sampling (bootstrap): Boolean values to toggle sampling with or without replacement.

The parameter ranges were carefully constructed to balance fine-grained sampling in smaller ranges and coarse exploration of broader intervals, accounting for dataset dimensionality and size. Using “scikit-learn”’s “RandomizedSearchCV” function, 5000 random parameter combinations were evaluated with 10-fold cross-validation on the training data, optimizing the specified scoring metric (default: accuracy). Parallel processing and verbosity were enabled for efficiency and transparency. The hyperparameter set achieving the best mean cross-validation score was selected as optimal. Comprehensive results, including all tested parameter combinations and corresponding performance metrics, were saved for transparency and reproducibility. The final set of optimised hyperparameters included the number of trees, maximum tree depth, minimum samples to split, minimum samples per leaf, maximum features per split, and bootstrap setting. These parameters informed subsequent model training and evaluation steps.

### Model training and evaluation

The model was trained using 80% of the data and evaluated using multiple performance metrics and visualisations to ensure a comprehensive assessment. A *k*-fold cross-validation approach was used to assess model generalisability. The dataset was partitioned into *k*=10 subsets; iteratively, one subset was held out as the test set while the model was trained on the remaining subsets. Accuracy scores were computed for each fold, and summary statistics (mean and standard deviation) were reported. Cross-validation predictions were aggregated to generate an overall confusion matrix. For binary classification, Receiver Operating Characteristic (ROC) and Precision-Recall (PR) curves were computed. The ROC curve was constructed by plotting true positive rate (sensitivity) against false positive rate (1 - specificity) at varying classification thresholds. The area under the ROC curve (AUC) served as a measure of classifier discrimination. PR curves were similarly generated by plotting precision against recall. For multi-class problems, one-vs-rest encoding was employed to generate ROC and PR curves for each class independently. Class-specific AUC values were calculated, and curves were visualised collectively for comparative analysis. Confusion matrices were generated to illustrate the number of true positive, true negative, false positive, and false negative predictions, providing insights into classification errors. Feature importance scores derived from the Random Forest model were analysed to identify the most influential variables contributing to classification performance. The top-N features were visualised to aid interpretability. All plots (ROC, PR, confusion matrix, feature importance) were saved as image files, and numerical data underlying these plots were exported as CSV files to facilitate reproducibility and further analysis.

### Canonical correlation analysis

CCA identifies linear combinations (*i.e.* canonical correlations) between two multivariate vectors (*i.e.* canonical variables) that are maximally correlated. Data for this analysis consisted of quantitative lipoprotein measurements alongside class labels indicating lipoprotein phenotypes (LP classes 1 to 3 from the previously identified clusters and an additional class LP4 with ∼130 lipoprotein phenotypes from healthy individuals). Data was imported into Python (v3.11.13) using the ‘pandas’ library. Class labels were encoded as integers for analysis. Predictor variables (X) included all quantitative features, while the outcome (Y) was defined as the one-hot encoded matrix of class labels. CCA was conducted to identify linear combinations of lipoprotein variables (X) and LP class variables (Y) that are maximally correlated.

Let *X* ∈ *R*^*n*.*p*^ and *Y* ∈ *R*^*n*.*q*^ denote the standardised data matrices for predictors and outcomes, respectively, with *n* observations, *p* number of predictor variables, and *q* number of outcome variables.

CCA seeks weight vectors *a* ∈ *R*^*p*^ and *b* ∈ *R*^*q*^ such that:

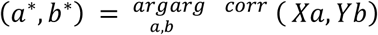

subject to the constraints that:

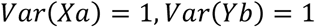

This optimisation yields pairs of canonical variates, *u* = *Xa* and *v* = *Yb*, which are orthogonal across pairs. The analysis was implemented using the CCA class from ‘scikit-learn’ (Pedregosa et al., 2011), with the number of components determined based on canonical correlation magnitudes and visual inspection of scree plots.

The canonical variate projections were computed via:

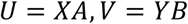

where *A* and *B* are matrices of canonical weights.

Canonical correlations *ρj* for each variate pair *j* were computed as:

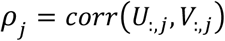

To identify the lipoprotein variables contributing most strongly to the canonical dimensions, canonical loadings were extracted from the coefficients matrix *A*. Variables were ranked by the absolute magnitude of their loadings for each canonical variate. For reporting, the top 20 features by absolute loading were retained for each canonical component, and the top 1% of features were exported for potential downstream analyses.

#### LPq score computation

A continuous severity score, LPq, was constructed from the first canonical variate as follows. For each observation *i*:

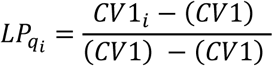

Prior to scaling, the sign of CV1 was examined. The direction of CV1 implied higher scores in healthy classes and lower scores in severe classes, therefore the sign was inverted such that higher LPq values consistently reflected more severe phenotypes. This ensured interpretability and comparability across analyses.

#### LPq using multiple canonical variates

Since CV1 primarily separated LP4, LP3 and LP2/LP1, while CV2 primarily separated LP4/LP3/LP2 and LP1, a weighted composite LPq score was constructed. Canonical variates were first standardised:

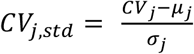

where *μj* and *σj* are the mean and standard deviation of the *j-th* canonical variate.

The weighted raw LPq score was then computed as:

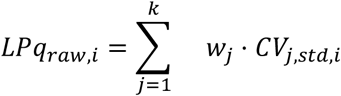

where weights *wj* were proportional to the canonical correlations:

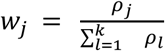

To transform the raw scores to the [0, 1] interval, the scaled LPq was calculated as:

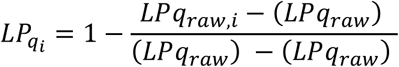

The inversion ensured that higher LPq values uniformly corresponded to greater severity across both single- and multi-variate analyses.

Exploratory and confirmatory data visualisations were generated in Python using ‘matplotlib’ and ‘seaborn’ libraries. Scatter plots, boxplots, violin plots, and histograms were used to assess the distribution of canonical variates and LPq scores across classes. Class-specific colour palettes were created based on discrete mappings of the “viridis” colormap, ensuring consistency across plots. Histograms stratified by class were plotted to visualise the distributional differences in LPq scores.

### Random Forest regression models

A Random Forest regression approach was employed to model the relationship between lipoprotein profiles and create a continuous severity score. The model pipeline included preprocessing steps to accommodate mixed data types and hyperparameter optimisation via Bayesian search, providing robust predictive performance and interpretability. The predictor variables were categorised into numeric and categorical features. Numeric features were standardised using z-score normalisation (mean = 0, standard deviation = 1) to ensure uniform scaling, while categorical features were encoded using one-hot encoding with unknown categories ignored during transformation. This preprocessing was implemented via a “ColumnTransformer” to integrate both scaling and encoding within our unified pipeline. A maximum of 30 iterations (n_iter=30) was performed in the Bayesian optimisation to identify the best hyperparameter configuration based on the coefficient of determination (R²) score.

## Data Availability

Data sets underlying our results have already been published in the previous papers.

https://www.combat.ox.ac.uk/

https://proteomecentral.proteomexchange.org/cgi/GetDataset?ID=PXD039875

## Declarations

## Funding

This work was funded in whole, or in part, by the Wellcome Trust Investigator Award (204969/Z/16/Z) (J.C.K.) and Wellcome Trust core funding to the Wellcome Sanger Institute (Grant numbers 206194 and 108413/A/15/D) and to the Centre for Human Genetics (090532/Z/09/Z); the Medical Research Council (MR/V002503/1) (J.C.K. and E.E.D.); the National Institute for Health Research (NIHR) Oxford Biomedical Research Centre (BRC) (J.C.K.); the Chinese Academy of Medical Sciences Innovation 537 Fund for Medical Science (2018-I2M-2-002) (PZ, JCK); and the NHS Genomic Medicine Service Genomic Network of Excellence in Severe Presentations of Infectious Disease (S.T.).

## Competing interests

M.N. reports consulting honoraria and a research grant to his institution from Baxter; Congress fee from Pfizer. J.C.K. reports a grant to his institution from the Danaher Beacon Programme for work on RNA biomarker point-of-care test development in sepsis.

## Acknowledgements

We thank all the patients, patient families, nurses, and clinicians who participated in the UK Genomic Advances in Sepsis (GAinS) and Sepsis Immunomics studies. MN wishes to thank the “Fondation Clément-Drevon” for their support.

## Availability of data and materials

Data sets underlying our results have already been published in the previous papers. All the scripts used to build and train the classification models, to do the canonical correlation analysis, to build and train the regression models, are available on GitHub at https://github.com/Sookie-S/Classifiers-for-risk-stratification-based-on-lipoproteins-proteome-alterations-in-sepsis. The functions that built the models and performed the hyperparameter optimisation for the classification and regression models are available as a python package that can be installed using “python pip install RFmodule”.

## Notes

### Author Declarations

This study consisted of a retrospective analysis of previously published prospective cohorts. Ethics approval was granted nationally and locally for individual participating centers, and we obtained informed consent from the patient or their legal representative (Scotland A Research Ethics Committee reference number 05/MRE00/38, Berkshire Research Ethics Committee (08/H0505/78), South Central Oxford REC C (19/SC/0296 and 06/Q1605/55).

