## Supplementary figures and images for "Inter-individual variability in lipoprotein proteomics reveals distinct patient clusters informative for disease pathogenesis and severity"

### Supplementary Figure 1

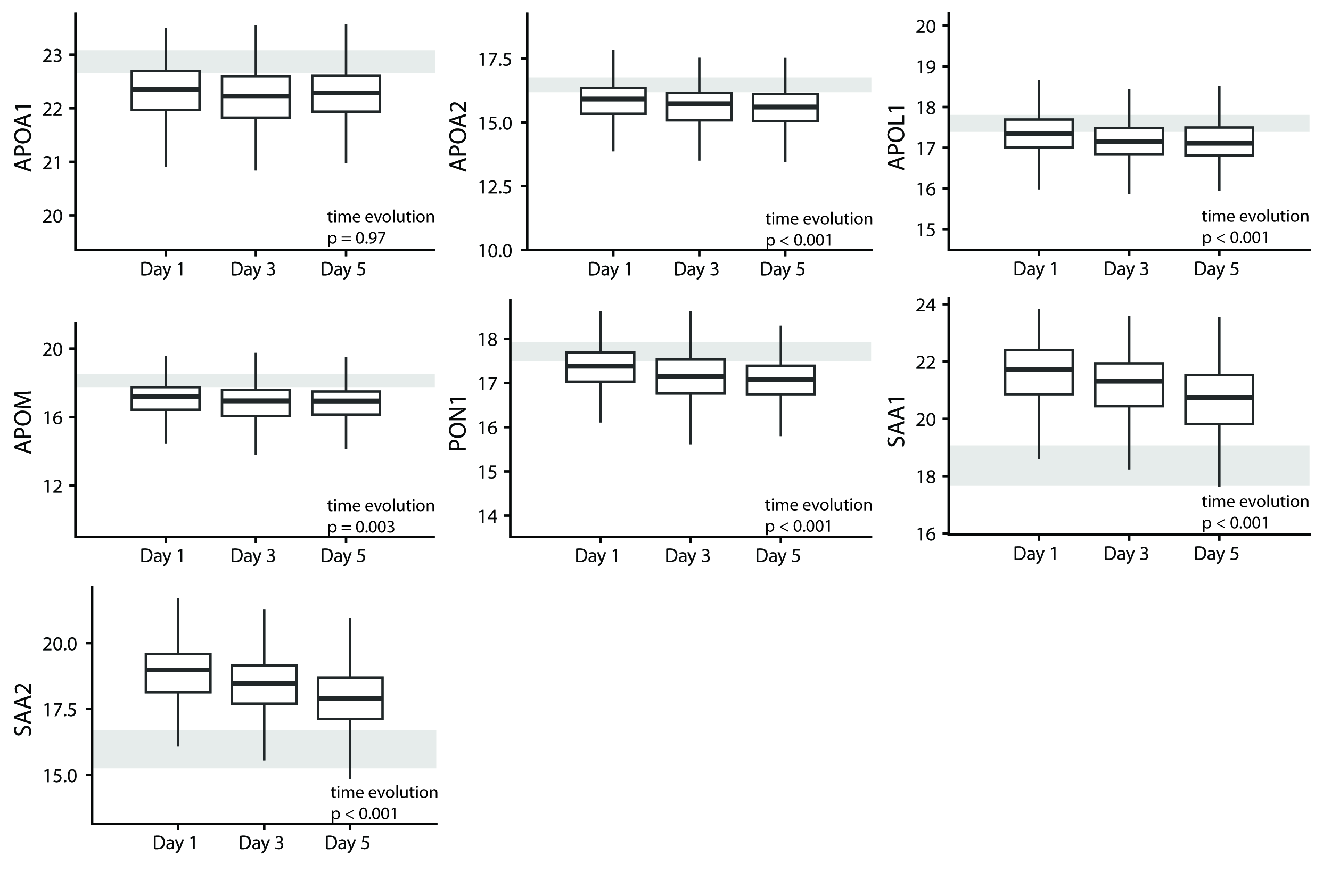

### Supplementary Figure 2

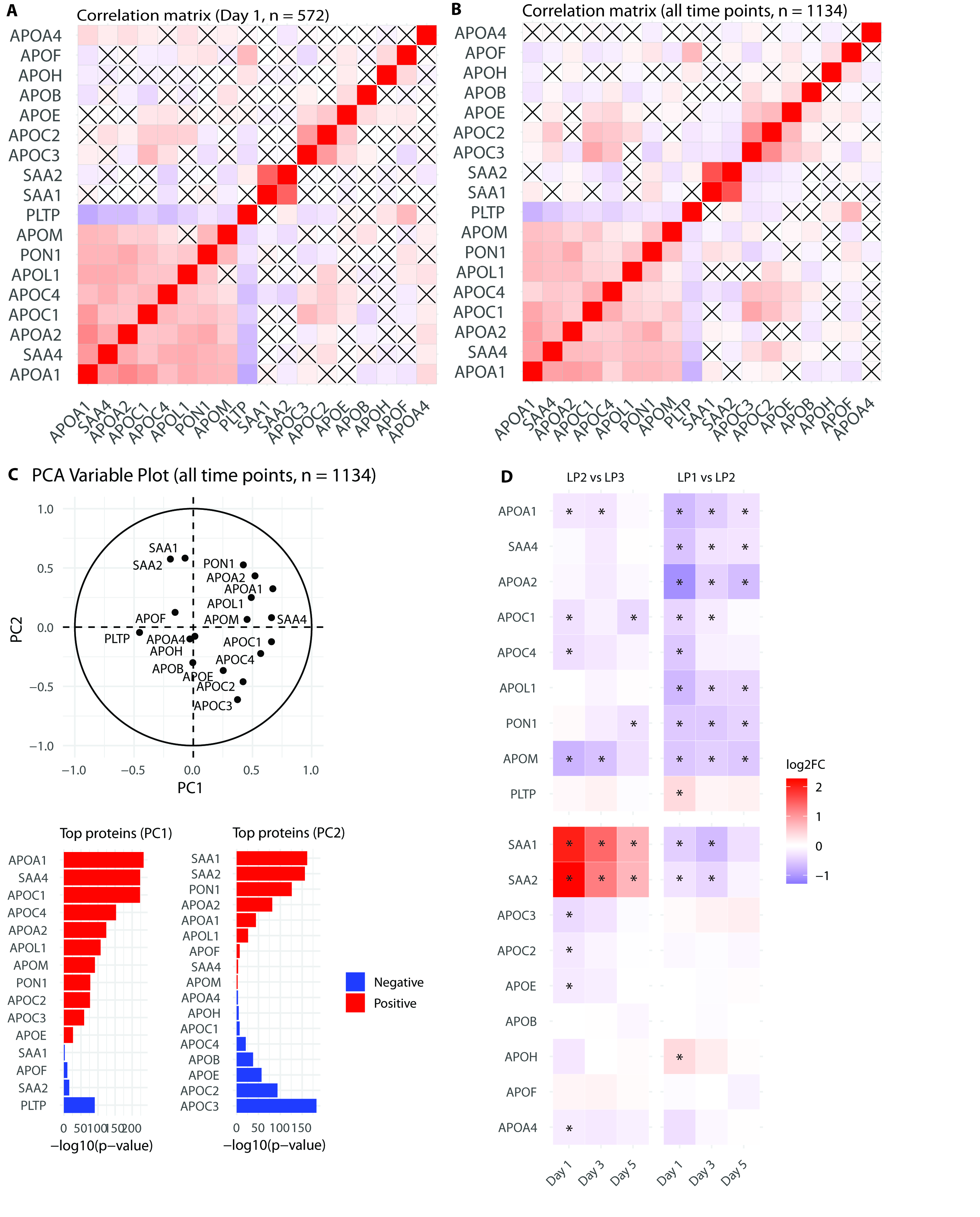

### Supplementary Figure 3

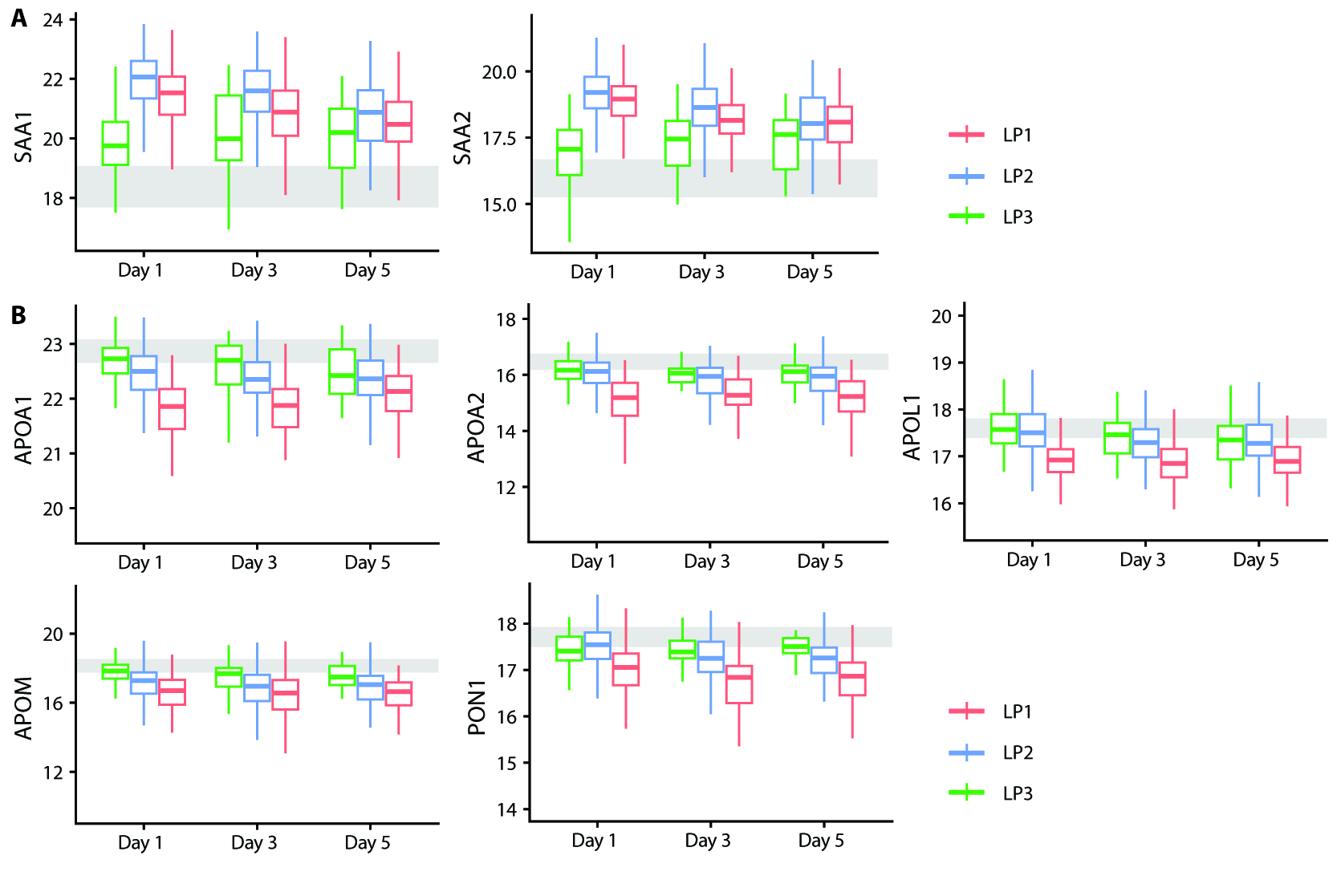

### Supplementary Figure 4

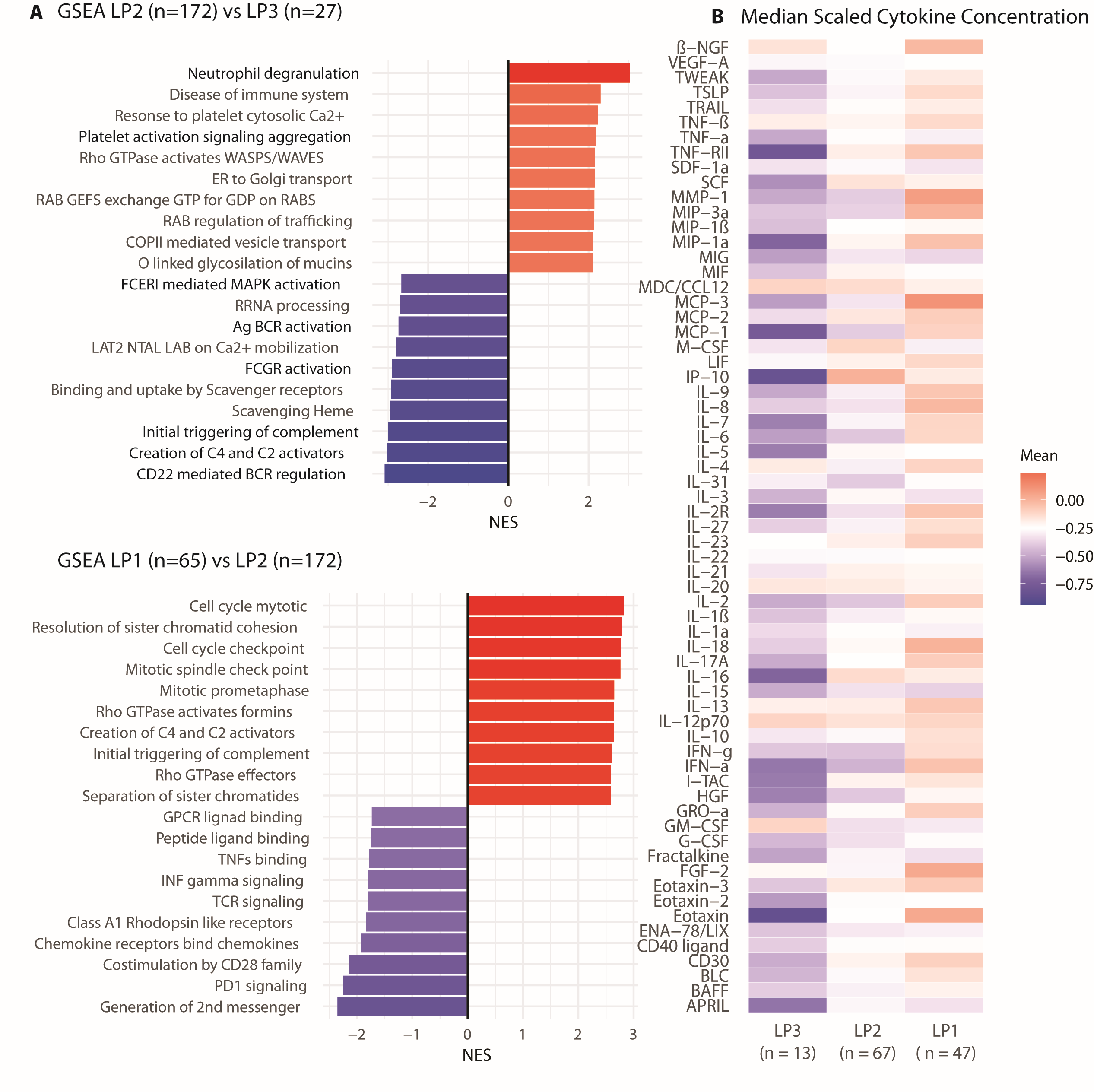

### Supplementary Figure 5

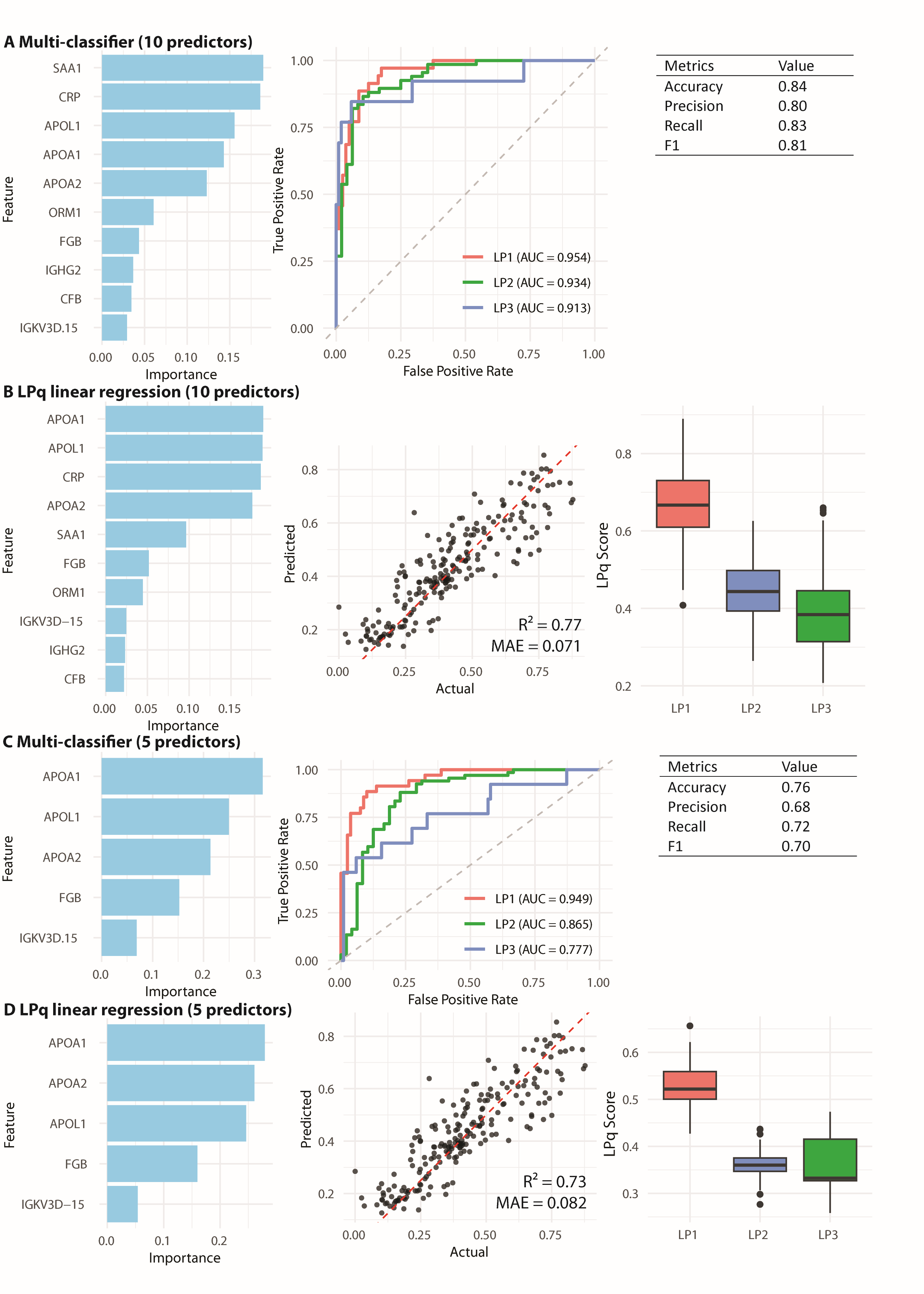

### Supplementary Figure 6

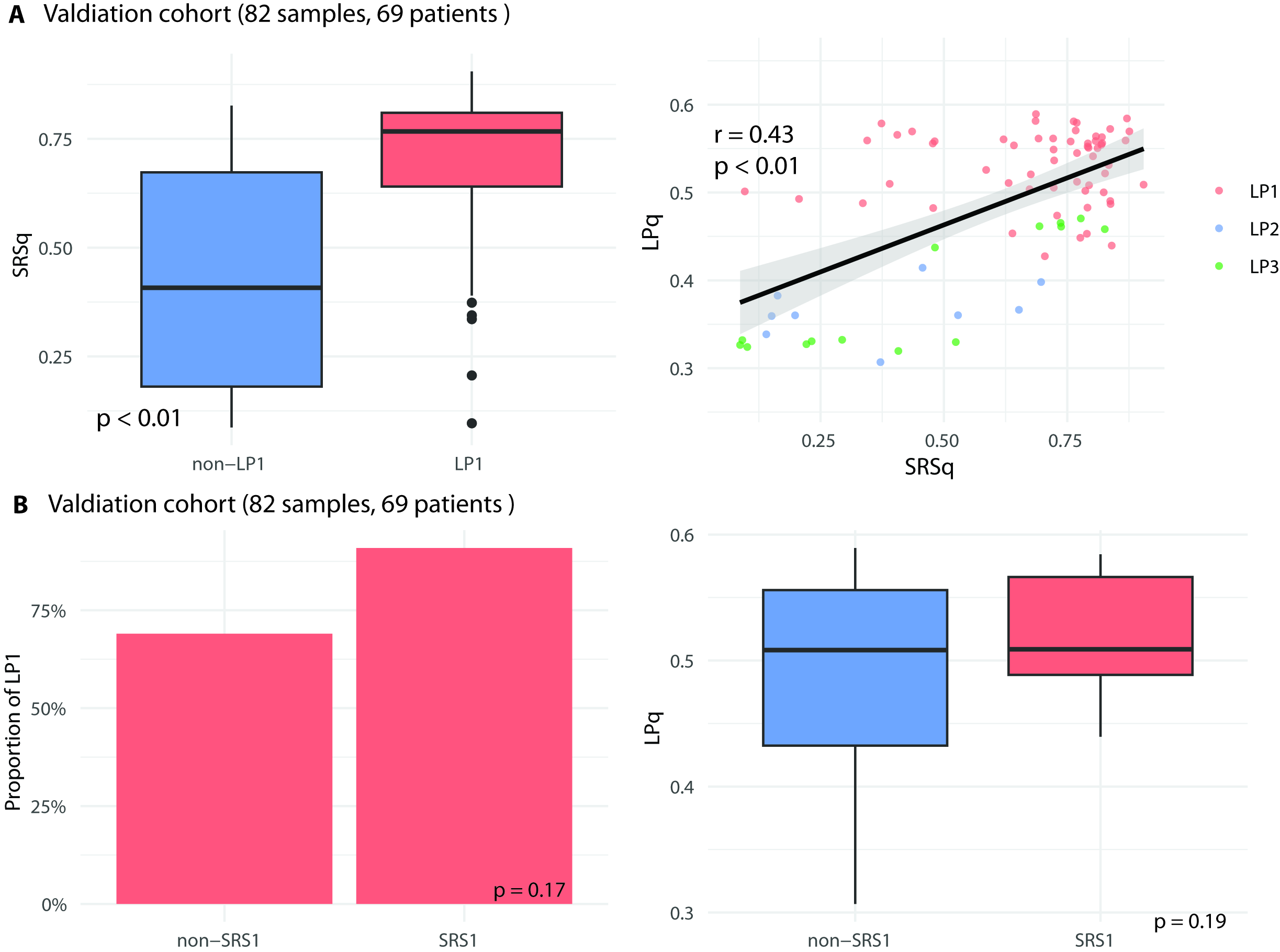
