## Supplementary Material 1 for "Inter-individual variability in lipoprotein proteomics reveals distinct patient clusters informative for disease pathogenesis and severity"

**Alterations of lipoproteins proteome are associated with immune dysregulation severity and mortality in sepsis : a cohort study**

Supplementary material 1

Methods for clustering

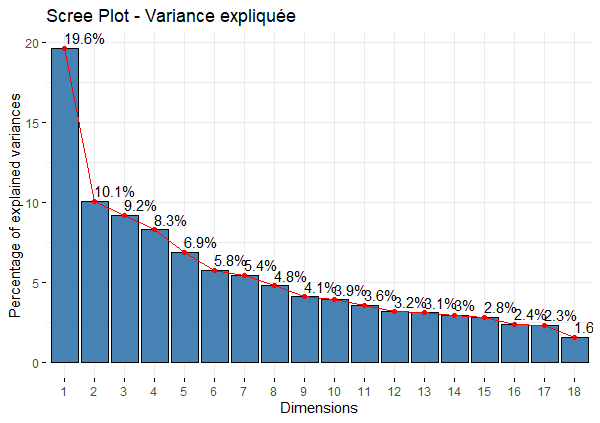

Figure 1. Scree plot showing the percentage of variance explained by each component.

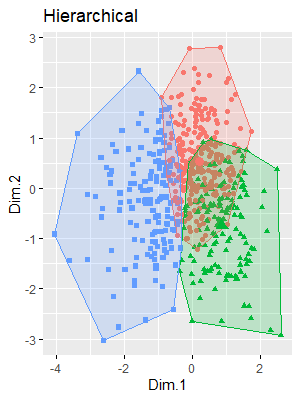

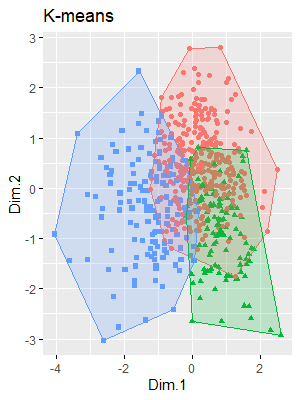

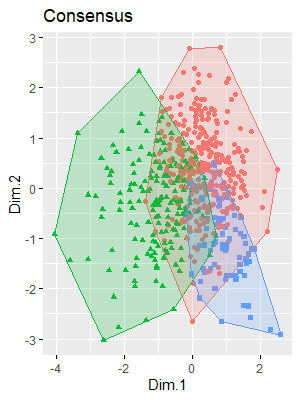

Figure 2. Cluster separation on the 2 first components according to the method of clustering

|  | Silhouette | Dunn | Calinski_Harabasz | Davies_Bouldin |
| --- | --- | --- | --- | --- |
| Hierarchical | **0.113** | 0.129 | 63.52 | **2.28** |
| K-means | 0.107 | 0.118 | **80.32** | 2.31 |
| Consensus | 0.106 | **0.130** | 68.49 | 2.29 |

Table 1. Parameters used for clustering method selection

|  | Number of patients | Proportion (%) |
| --- | --- | --- |
| LP1 | 172 | 30.1 |
| LP2 | 333 | 58.2 |
| LP3 | 67 | 11.7 |

Table 2. Number and proportion of patients in each cluster

**GAinS Investigators** The following GainS investigators, listed alphabetically by institution, were involved in patient recruitment, sample collection, or sample processing:

Jenni Addison^1^, Helen Galley^1^, Sally Hall^1^, Sian Roughton^1^, Jane Taylor^1^, Heather Tennant^1^, Nigel Webster^1^, Achyut Guleri^2^, Natalia Waddington^2^, Dilshan Arawwawala^3^, John Durcan^3^, Christine Mitchell-Inwang^3^, Alasdair Short^3^, Susan Smolen^3^, Karen Swan^3^, Sarah Williams^3^, Emily Errington^4^, Tony Gordon^4^, Maie Templeton^4^, Marie McCauley^5^, Pyda Venatesh^5^, Geraldine Ward^5^, Simon Baudouin^6,23^, Sally Grier^7^, Elaine Hall^7^, Charley Higham^6^, Jasmeet Soar^7^, Stephen Brett^8^, David Kitson^8^, Juan Moreno^8^, Laura Mountford^8^, Robert Wilson^8^, Peter Hall^9^, Jackie Hewlett^9^, Stuart McKechnie^10,11^, Roser Faras-Arraya^11^, Christopher Garrard^11^, Paula Hutton^11^, Julian Millo^11^, Penny Parsons^11^, Alex Smiths^11^, Duncan Young^11^, Parizade Raymode^12^, Jasmeet Soar^12^, Prem Andreou^13^, Sarah Bowrey^13^, Dawn Hales^13^, Sandra Kazembe^13^, Natalie Rich^13^, Emma Roberts^13^, Jonathan Thompson^13^, Simon Fletcher^14^, Georgina Glister^14^, Melissa Rosbergen^14^, Jeronimo Moreno Cuesta^15^, Julian Bion^16^, Ronald Carrera^16^, Sarah Lees^16^, Joanne Millar^16^, Natalie Mitchell^16^, Annette Nilson^16^, Elsa Jane Perry^16^, Sebastian Ruel^16^, Jude Wilde^16^, Heather Willis^16^, Jane Atkinson^17^, Abby Brown^17^, Nicola Jacques^17^, Atul Kapila^17^, Heather Prowse^17^, Martin Bland^18^, Lynne Bullock^18^, Donna Harrison^18^, Anton Krige^18^, Gary Mills^19,20^, John Humphreys^19,20^, Kelsey Armitage^19,20^, Shond Laha^21^, Jacqueline Baldwin^21^, Angela Walsh^21^, Nicola Doherty^21^, Stephen Drage^22^, Laura Ortiz-Ruiz de Gordoa^22^, Sarah Lowes^22^, Charley Higham^23^, Helen Walsh^23^, Verity Calder^23^, Catherine Swan^23^, Heather Payne^23^, David Higgins^24^, Sarah Andrews^24^, Sarah Mappleback^24^, Charles Hinds^25,32^, D Watson^26,27^, Eleanor McLees^26,27^, Alice Purdy^26,27^, Martin Stotz^28^, Adaeze Ochelli-Okpue^28^, Stephen Bonner^29^, Iain Whitehead^29^, Keith Hugil^29^, Victoria Goodridge^29^, Louisa Cawthor^29^, Martin Kuper^30^, Sheik Pahary^30^, Geoffrey Bellingan^31^, Richard Marshall^31^, Hugh Montgomery^31^, Jung Hyun Ryu^31^, Georgia Bercades^31^, Susan Boluda^31^, Andrew Bentley^32^, Katie Mccalman^32^, Fiona Jefferies^32^, Alice Allcock^33^, Katie Burnham^33^, Emma Davenport^33^, Cyndi Geoghegan^33^, Julian Knight^33^, Narelle Maugeri^33^, Yuxin Mi^33^, and Jayachandran Radhakrishnan^33^.

1.Aberdeen Royal Infirmary, Aberdeen AB25 2ZN, UK.

2.Blackpool Victoria Hospital, Blackpool FY3 8NR, UK.

3.Broomfield Hospital, Chelmsford CM1 7ET, UK.

4.Charing Cross Hospital, London W6 8RF, UK.

5.Coventry and Warwickshire University Hospital, Coventry CV2 2DX, UK.

6.Freeman Hospital, Newcastle upon Tyne NE7 7DN, UK.

7.Frenchay Hospital, Bristol, UK and Southmead Hospital, Bristol BS16 1JE, UK. 8.Hammersmith Hospital, London W12 0HS, UK.

9.Huddersfield Royal Infirmary, Huddersfield HD3 3EA, UK.

10.Oxford University Hospitals NHS Foundation Trust, Oxford

11.John Radcliffe Hospital, Headington, Oxford OX3 9DU, UK.

12.Kettering General Hospital, Kettering NN16 8UZ, UK.

13.Leicester Royal Infirmary, Leicester LE1 5WW, UK.

14.Norfolk and Norwich University Hospital, Norwich NR4 7UY, UK.

15.North Middlesex Hospital, London N18 1QX, UK.

16.Queen Elizabeth Hospital, Birmingham B15 2GW, UK.

17.Royal Berkshire Hospital, Reading RG1 5AN, UK.

18.Royal Blackburn Hospital, Blackburn BB2 3HH, UK. 2

19.Royal Hallamshire Hospital, Sheffield S10 2JF, UK.

20.Northern General Hospital, Sheffield S5 7AU, UK.

21.Royal Preston Hospital, Preston PR2 9HT, UK.

22.Royal Sussex County Hospital, Brighton BN2 5BE, UK.

23.Royal Victoria Infirmary, Newcastle upon Tyne NE1 4LP, UK.

24.Southend Hospital, Westcliff-on-Sea SS0 0RY, UK.

25.Centre for Translational Medicine & Therapeutics, William Harvey Research Institute, Faculty of Medicine & Dentistry, Queen Mary University of London, London, UK

26.St Bartholomew’s Hospital, London EC1A 7BE, UK.

27.Royal London Hospital, London E1 1FR, UK.

28.St Mary’s Hospital, London W2 1NY, UK.

29.James Cook University Hospital, Middlesbrough TS4 3BW, UK.

30.Whittington Hospital, London N19 5NF, UK.

31.University College London Hospital, UCLH, London NW1 2BU, UK.

32.Wythenshawe Hospital, Manchester M23 9LT, UK.

33.Centre for Human Genetics, University of Oxford, Oxford, UK
